# Discovery of Sexual Dimorphism in the Serum Metabolome of Parkinson’s Disease Patients Harboring Rare Genetic Variants with Uncertain Pathogenicity

**DOI:** 10.1101/2025.11.20.25340670

**Authors:** Carmen Marino, Federica Carrillo, Tommaso Nuzzo, Marcello Serra, Isar Yahyavi, Manuela Grimaldi, Sara Pietracupa, Nicola Modugno, Francesco Errico, Anna Maria D’Ursi, Teresa Esposito, Alessandro Usiello

## Abstract

Recent findings show that sex and genetic background impact serum metabolic profiles in idiopathic Parkinson’s disease (iPD) and carriers of pathogenic mutations (gPD). However, the metabolic consequences in patients harboring rare variants of uncertain pathogenic significance remain poorly understood. Here, we combined ^1^H-Nuclear Magnetic Resonance (NMR) metabolomics with high-performance liquid chromatography (HPLC) to characterize serum metabolic features in 336 clinically and genetically defined PD patients (iPD, gPD, and rare-variant PD [rvPD]) and 137 sex-matched healthy controls (HCs). Notably, our findings revealed significant differences in blood metabolome profiles between individuals with rvPD and controls. Additionally, sex-stratified analyses disclosed a marked effect of sex on circulating metabolic profiles between cases and controls. Specifically, male rvPD patients exhibited more pronounced disruptions in amino-acid metabolism, whereas females displayed lipid-related pathway alterations. Interestingly, multivariate analysis of NMR data showed no significant differences among patients with rvPD, iPD, and gPD, indicating shared systemic biochemical alterations across subtypes. HPLC analysis of serum D- and L-amino acids involved in glutamatergic transmission supports NMR-based findings, highlighting a reduction in L-Glu levels only in male rvPD patients compared to matched controls. In conclusion, our findings demonstrate that sex differences shape systemic metabolomic features in PD patients carrying rare genetic variants.

## Introduction

Parkinson’s disease (PD) is a neurodegenerative disorder whose incidence increases with age, characterised by the progressive loss of dopaminergic neurons in the substantia nigra pars compacta (SNpc), and the formation of intracellular Lewy bodies composed primarily of α-synuclein aggregates ^1–5^. PD presents with motor and non-motor symptoms, often preceded by a prodromal phase with non-motor features such as hyposmia, constipation, and sleep disturbances ^6^. Despite the well-characterised clinical features, the etiological factors underlying PD onset remain under investigation, with genetic factors estimated to account for approximately 25% of the overall risk ^7^. Indeed, with the advancement of genome-wide association studies (GWAS) in large cohorts of PD patients and controls, more than 100 loci have been identified as risk factors for PD, while studies on familial cases using whole-exome and whole-genome sequencing (WES/WGS) have led to the identification of over 20 monogenic forms of the disease ^7–11^.

Monogenic variants—either highly penetrant or of reduced penetrance—in genes such as *SNCA*, *PARK2*, *PARK7*, *LRRK2*, *VPS35*, and *PINK1* have been causally linked to PD and together account for approximately 5–10 % of all cases ^10,12^. However, it is known that the spectrum of genetic variants underlying PD aetiology could range from rare variants with a considerable effect (fully or highly penetrant mutations in single genes) to genetic variants exerting only modest effects, which are relatively common in the general population. Furthermore, rare variants in Mendelian PD genes can also act as risk factors for late-onset, sporadic PD ^13^. Recent studies have highlighted the extreme genetic heterogeneity among PD patients, in terms of the number of rare variants across different genes associated with PD. To date, only a small fraction of these rare variants has been functionally characterized and classified as pathogenic for PD, while most remain uncharacterized. Segregation studies in informative PD families have often not been sufficient to confirm their pathogenicity. In this complex context, a polygenic rare variant load model of inheritance has been suggested in PD patients ^14–16^. This model proposes that the co-inheritance of multiple rare variants in Mendelian genes may increase the risk of PD in a non-Mendelian fashion and influence both disease onset and clinical presentation ^15,16^. Nonetheless, even a single rare variant in PD-related genes, although with lower statistical power, can be predictive of the disease ^15,16^

In recent years, clinical, imaging, and genomic biomarkers have been extensively investigated to improve PD diagnosis and to monitor disease progression ^17–22^. In parallel, a growing body of studies has examined the blood metabolome in patients with PD ^23–31^. Metabolic alterations identified in different biological specimens have shown potential for discriminating patients with PD from healthy controls (HCs) and for providing insights into underlying pathophysiological mechanisms ^32^. Taken together these findings support the view of PD as a complex multiorgan and multisystem disorder involving peripheral metabolic dysfunction rather than a pathological condition restricted to the basal ganglia ^33,34^.

In line with this notion, circulating metabolomic alterations have been consistently reported in male and female patients with idiopathic PD (iPD) and in carriers of pathogenic mutations (genetic PD), particularly involving amino acid, lipid, antioxidant molecules and energy-related pathways ^31,35–44^. However, studies specifically examining patients harboring rare, deleterious variants of uncertain pathogenicity remain extremely limited. Moreover, the influence of sex on metabolic profiles in this subgroup is currently unknown, despite our previous work demonstrating that sex is a major determinant of serum metabolomic signatures in PD, with male patients exhibiting more pronounced amino acids alterations than females across both iPD and gPD cohorts ^25,26,45–48^.

To gain insight into this issue, in the present work we conducted an untargeted ¹H-Nuclear Magnetic Resonance (NMR)-based metabolomic analysis of blood serum from a cohort of clinically and genetically well-characterized PD patients carrying at least one rare variant not yet confirmed as pathogenic—hereafter referred to as rare variant PD patients (rvPD)—and compared their profiles with those of sex-matched HCs and patients with either iPD or gPD. Furthermore, we performed High-Performance Liquid Chromatography (HPLC) analysis on the same samples to measure circulating levels of amino acids involved in glutamatergic NMDA receptor (NMDAR) signalling and their precursors, which have been previously linked to PD pathophysiology and L-DOPA treatment ^49–52^. Additionally, in rvPD patients, we also evaluated potential correlations between blood D- and L-amino acid levels and demographic and clinical variables including: (i) age; (ii) disease duration; (iii) age at onset; (iv) L-DOPA equivalent daily dose (LEDD); (v) Movement Disorder Society-Unified Parkinson’s Disease Rating Scale (MDS-UPDRS) scores. Finally, we investigated the genetic impact of a large panel of variants in genes encoding NMDAR subunits and key enzymes involved in glycine and serine metabolism to disclose their potential role as context-dependent modifiers.

## Results

### Clinical and demographic characteristics of patients with Parkinson’s disease and healthy control individuals

137 HCs and 336 PD patients were included in this case-control observational study. Demographic and clinical features of participants are reported in **Table 1 and Table 2**. In the PD cohort, 121 patients were classified as idiopathic (iPD; 65 male/56 female; **Table 1**), 124 as genetic PD carrying pathogenic variants (gPD, 64 male/60 female; **Table 1**) and 91 exhibited rare variants not yet confirmed as pathogenic (rvPD; 44 male/47 female; **Table 1**). The complete list of mutations/variants identified in gPD and rvPD patients is reported in **Table S1**. The age of HCs ranged from 43 to 90 years, whereas the age of iPD, gPD, and rvPD patients ranged from 48 to 82 years, from 41 to 85, and from 40 to 79 years, respectively. Both iPD, gPD, and rvPD patients were significantly older than HCs (median: HC = 62 years; iPD, gPD = 68 years; rvPD = 67 years; **Table 1**), whereas sex distribution did not differ among groups (**Table 1**). Moreover, sex distribution, age, age at onset, disease duration, LEDD, and MDS-UPDRS scores were comparable across iPD, gPD, and rvPD patient groups (**Table 1**). Consistently, no significant differences in demographic or clinical parameters were observed among these groups following sex stratification. Although the Kruskal–Wallis analysis evidenced an overall difference in disease duration across groups in females (median: HC = 5 years; iPD, gPD = 7 years; rvPD = 5 years; p = 0.033; **Table S2**), this was not supported by post-hoc analysis (p = 0.067).

**Table 1.**
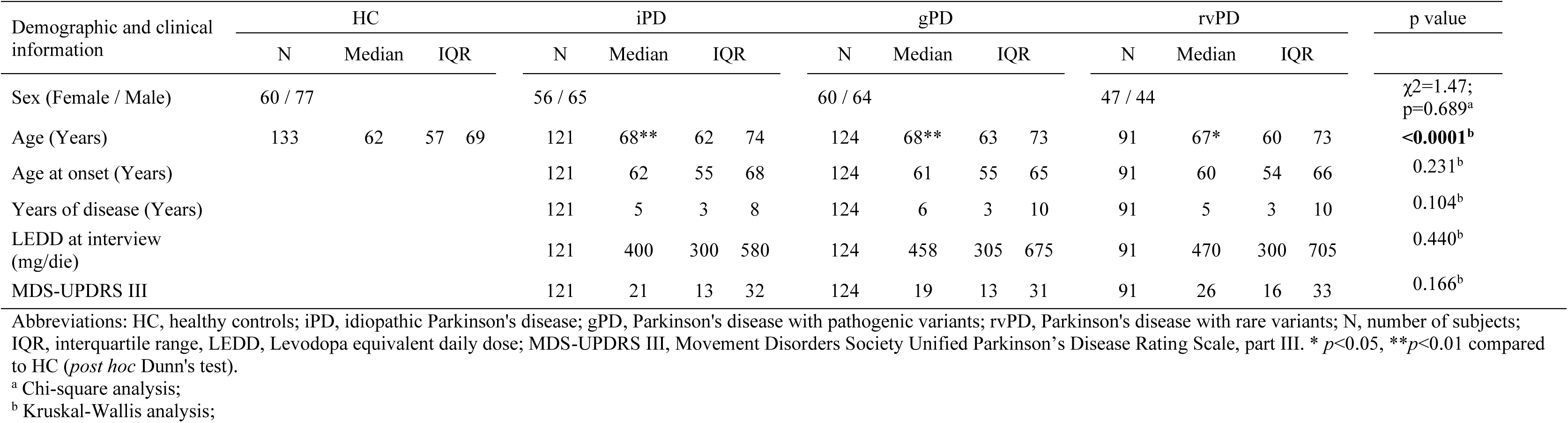
Demographic and clinical characteristics of PD patients and HC enrolled in the serum collection.

| Demographic and clinical information | HC |  |  | iPD |  |  |  | gPD |  |  |  | rvPD |  |  |  | p value |
| --- | --- | --- | --- | --- | --- | --- | --- | --- | --- | --- | --- | --- | --- | --- | --- | --- |
|  | N | Median | IQR | N | Median | IQR |  | N | Median | IQR |  | N | Median | IQR |  |  |
| Sex (Female / Male) | 60 / 77 | | | 56 / 65 | | | | 60 / 64 | | | | 47 / 44 | | | | $\chi^2=1.47$;<br>p=0.689<sup>a</sup> |
| Age (Years) | 133 | 62 | 57 69 | 121 | 68** | 62 74 |  | 124 | 68** | 63 73 |  | 91 | 67* | 60 73 |  | <0.0001<sup>b</sup> |
| Age at onset (Years) |  |  |  | 121 | 62 | 55 68 |  | 124 | 61 | 55 65 |  | 91 | 60 | 54 66 |  | 0.231<sup>b</sup> |
| Years of disease (Years) |  |  |  | 121 | 5 | 3 8 |  | 124 | 6 | 3 10 |  | 91 | 5 | 3 10 |  | 0.104<sup>b</sup> |
| LEDD at interview (mg/die) |  |  |  | 121 | 400 | 300 580 |  | 124 | 458 | 305 675 |  | 91 | 470 | 300 705 |  | 0.440<sup>b</sup> |
| MDS-UPDRS III |  |  |  | 121 | 21 | 13 32 |  | 124 | 19 | 13 31 |  | 91 | 26 | 16 33 |  | 0.166<sup>b</sup> |

Abbreviations: HC, healthy controls; iPD, idiopathic Parkinson's disease; gPD, Parkinson's disease with pathogenic variants; rvPD, Parkinson's disease with rare variants; N, number of subjects; IQR, interquartile range, LEDD, Levodopa equivalent daily dose; MDS-UPDRS III, Movement Disorders Society Unified Parkinson's Disease Rating Scale, part III. \* $p<0.05$ , \*\* $p<0.01$ compared to HC (*post hoc* Dunn's test).
<sup>a</sup> Chi-square analysis;
<sup>b</sup> Kruskal-Wallis analysis;

**Table 2.** Concentrations of serum D- and L- amino acids levels in male and female PD patients carrying rare genetic variants and HC subjects.

| Sex | Amino acids | HC |  |  |  | rvPD |  |  |  | Mann-Whitney | ANCOVA<sup>a</sup> |  |
| --- | --- | --- | --- | --- | --- | --- | --- | --- | --- | --- | --- | --- |
|  |  | N | Median | IQR |  | N | Median | IQR |  | *p* value | F<sub>(1;114)</sub> | *p* value |
| Male | L-Asp (μM) | 77 | 12.1 | 8.8 | 16 | 44 | 10.7 | 8.2 | 14.6 | 0.127 |  |  |
|  | L-Glu (μM) | 77 | 31 | 21.9 | 43.1 | 44 | 23.7 | 17.5 | 35.6 | **0.026** | 4.035 | **0.047** |
|  | L-Asn (μM) | 77 | 22.6 | 15.2 | 34.2 | 44 | 18.3 | 10.8 | 27.4 | 0.074 |  |  |
|  | D-Ser (μM) | 77 | 2.8 | 2.4 | 3.2 | 44 | 2.8 | 2.1 | 3.1 | 0.537 |  |  |
|  | L-Ser (μM) | 77 | 74.9 | 52.9 | 106.4 | 44 | 58.3 | 42.6 | 83.9 | 0.077 |  |  |
|  | D-Ser/total Ser (%) | 77 | 3.5 | 2.4 | 4.7 | 44 | 3.9 | 2.6 | 5.6 | 0.242 |  |  |
|  | L-Gln (μM) | 77 | 182.2 | 136 | 238.9 | 44 | 157.7 | 107 | 195 | 0.168 |  |  |
|  | Gly (μM) | 77 | 152.4 | 108 | 201.1 | 44 | 123.9 | 94.1 | 150 | 0.051 |  |  |
|  | L-Gln/L-Glu | 77 | 6.1 | 4.4 | 7.2 | 44 | 6.5 | 5.1 | 8 | 0.093 |  |  |
| Female | L-Asp (μM) | 60 | 9.7 | 7 | 15 | 47 | 10.7 | 8.3 | 14.6 | 0.510 |  |  |
|  | L-Glu (μM) | 60 | 22.3 | 13 | 34.6 | 47 | 25.5 | 16.2 | 34.6 | 0.518 |  |  |
|  | L-Asn (μM) | 60 | 15.7 | 8.5 | 27.3 | 47 | 22 | 12.4 | 28.3 | 0.175 |  |  |
|  | D-Ser (μM) | 60 | 2.6 | 2 | 3.1 | 47 | 2.5 | 2.1 | 3.1 | 0.885 |  |  |
|  | L-Ser (μM) | 60 | 57.1 | 32 | 103.8 | 47 | 65 | 44.9 | 100 | 0.433 |  |  |
|  | D-Ser/total Ser (%) | 60 | 3.8 | 2.4 | 6.4 | 47 | 3.6 | 2.6 | 4.9 | 0.526 |  |  |
|  | L-Gln (μM) | 60 | 140.2 | 87.1 | 213.1 | 47 | 176.3 | 119 | 233 | 0.099 |  |  |
|  | Gly (μM) | 60 | 140.4 | 94.7 | 195.2 | 47 | 158.8 | 116 | 214 | 0.084 |  |  |
|  | L-Gln/L-Glu | 60 | 6.2 | 4.6 | 7.4 | 47 | 6.7 | 5.5 | 8.7 | 0.074 |  |  |

Abbreviations: HC, healthy controls; rvPD, Parkinson's disease with rare variants; N, number of subjects; IQR, interquartile range.
<sup>a</sup> adjusted for age and LEDD

### ¹H-NMR-based metabolomics in blood serum reveals distinct biochemical profiles in patients carrying rare genetic variants compared to healthy controls

First, we examined the serum metabolome profiles of rvPD patients (N = 91) and HCs (N = 137), regardless of sex differences.^1^H 1D NMR experiments using CPMG pulse sequence allowed the identification of 43 analytes using CHENOMX software (**Figure S1**). These metabolites were then quantified and subjected to statistical analysis using MetaboAnalyst 6.0 ^53^

Notably, the PLS-DA score plot revealed a significant separation in the serum metabolomic profiles between rvPD patients and HCs (Q^2^ values for PC1 0.71) (**Fig. 1a**). Specifically, VIP analysis revealed notable discriminatory power among different amino acids in clustering the metabolomic profiles of cases and controls. Accordingly, the blood metabolomic profile of patients with rvPD was distinct from that of HCs for L-glutamic acid (VIP: 4.17), L-glutamine (VIP: 1.76), valine (VIP: 1.44), and L-ornithine (VIP: 1.36). Furthermore, other molecules such as 3-hydroxybutyrate (VIP: 1.71), acetic acid (VIP: 1.64), pyruvic acid (VIP: 1.34), choline (VIP: 1.17) and malonate (VIP: 1.04) emerged as metabolites that differentiate the serum profile of rvPD patients from HCs (**Fig. 1b**). Robust volcano plot indicated a significant reduction in serum L-glutamic acid and pyruvic acid levels concomitant with an increase in L-glutamine levels in patients with rvPD in comparison to HCs (**Fig. 1c, Table S3**). Furthermore, the heatmap analysis showed elevated serum levels of L-ornithine in the rvPD group, along with higher concentrations of 3-hydroxybutyrate and isobutyric acid compared to HCs. Conversely, reduced levels of valine and acetic acid were observed in cases compared to controls (**Fig. 1e**).

**Figure 1.**
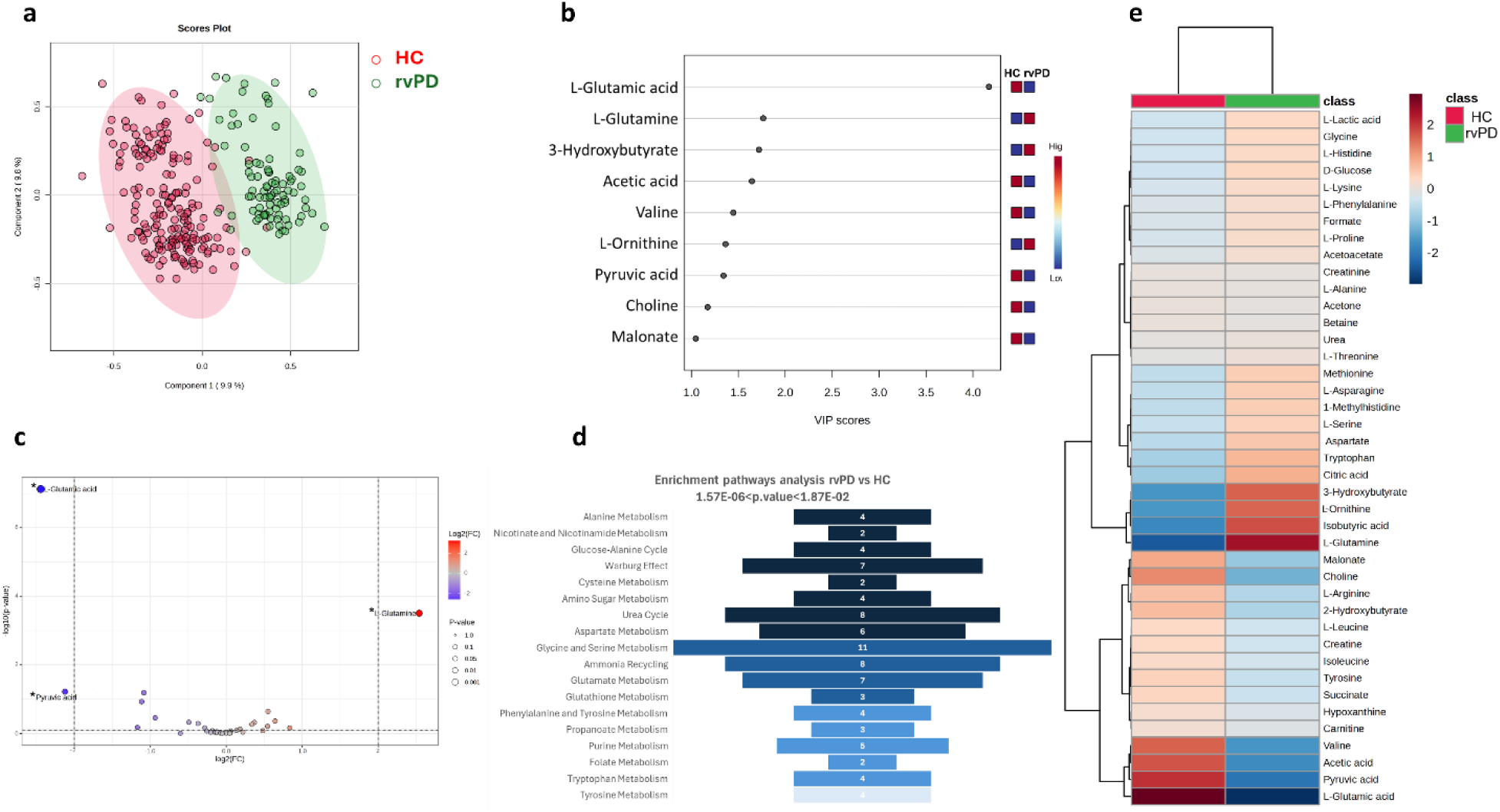
¹H-NMR metabolomics in blood serum uncovers unique biochemical signatures in patients with rare genetic variants compared to healthy controls. **a,** The Partial Least Squares Discriminant Analysis (PLS-DA) score plot displaying the percentage of variance attributed to the primary (PC1, x-axis) and secondary (PC2, y-axis) components of the model (PC1: 9.9%; PC2: 9.8%). The supervised model was developed based on the serum metabolite concentrations of 91 PD patients (green cluster) and 137 healthy controls (red cluster). The model underwent validation using a 10-fold cross-validation (CV) approach, reporting accuracy percentages for PC1 and PC2 of 0.94 and 0.96, respectively, along with Q² values of 0.71 and 0.75, respectively. **b,** Variable Importance Projection (VIP) plot reporting the metabolites responsible for cluster separations. Only metabolites with VIP > 1 were considered significant. **c,** Robust volcano plot displaying the upregulated (red) and downregulated (blue) metabolites in the pathogenic cluster serum. The fold-change value was set to 2, while the p-value threshold was set to <0.05. Metabolites significant for both fold-change and p-value from the t-test are labelled with an asterisk. **d,** Pathway enrichment analysis was performed on ¹H-NMR data related to serum metabolomic results of rvPD patients compared to healthy controls (HC). The bars represent the hits, i.e., the number of metabolites detected in the spectra and involved in the pathways. Pathways were considered statistically significant with hits > 1, p-value < 0.05, and p-values adjusted using the Holm–Bonferroni test (Holm p) and the False Discovery Rate (FDR) < 1. Darker colours represent more significant p-values. The range of significance is shown at the top. **e,** Heatmap illustrating altered metabolites in rvPD patients compared to HC. The colour of each section corresponds to the concentration value of each metabolite, calculated from a normalised concentration matrix (red, upregulated; blue, downregulated).

In line with a considerable reduction in glutamic acid, a pathway enrichment analysis using the Small Molecule Pathways Database (SMPDB) ^53^, revealed a significant dysregulation of various amino acid metabolic pathways in patients compared to controls. Thus, besides glutamate, in rvPD patients, we found significant alterations in the metabolism of alanine, cysteine, aspartate, glycine-serine, phenylalanine-tyrosine, and tryptophan, compared to HCs (**Fig. 1d, Table S4)**. Notably, the reduction of glutamate also influenced the glutathione biosynthesis (**Fig. 1d, Table S4)**. Untargeted NMR data analysis also revealed significant dysregulation of key pathways involved in bioenergetic homeostasis, including the glucose-alanine cycle, the Warburg effect, amino sugar metabolism and the nicotinate and nicotinamide metabolism (**Fig. 1d, Table S4**).

Additionally, abnormalities in metabolites implicated in the urea cycle and ammonia recycling, along with alterations in folate, propanoate and purine metabolism were observed in rvPD patients compared to HCs (**Fig. 1d, Table S4).**

In summary, our NMR findings highlight profound systemic disruptions in amino acid metabolism, antioxidant molecules biosynthesis, and bioenergetic pathways in patients harbouring rare variants compared with HCs, which align well with systemic metabolic alterations previously reported in PD pathophysiology ^28,31,46,54–59^.

### Sex differences impact serum metabolome profiles in PD patients harbouring rare genetic variants compared to healthy controls

Here, we investigated whether sex differences significantly modulate the serum metabolome features in PD patients carrying at least one rare genetic variant of uncertain pathogenicity, in comparison with sex-matched HCs.

Notably, score plots from the supervised PLS-DA analysis indicated a significant separation of the serum metabolomes between rvPD patients and HCs in both males (patients, n = 44; controls, n = 77) (**Fig. 2a**) and females (patients, n = 47; controls, n = 60) (**Fig. 3a**), as indicated by the PC1’s Q^2^ values of 0.73 and 0.64, respectively.

**Figure 2.**
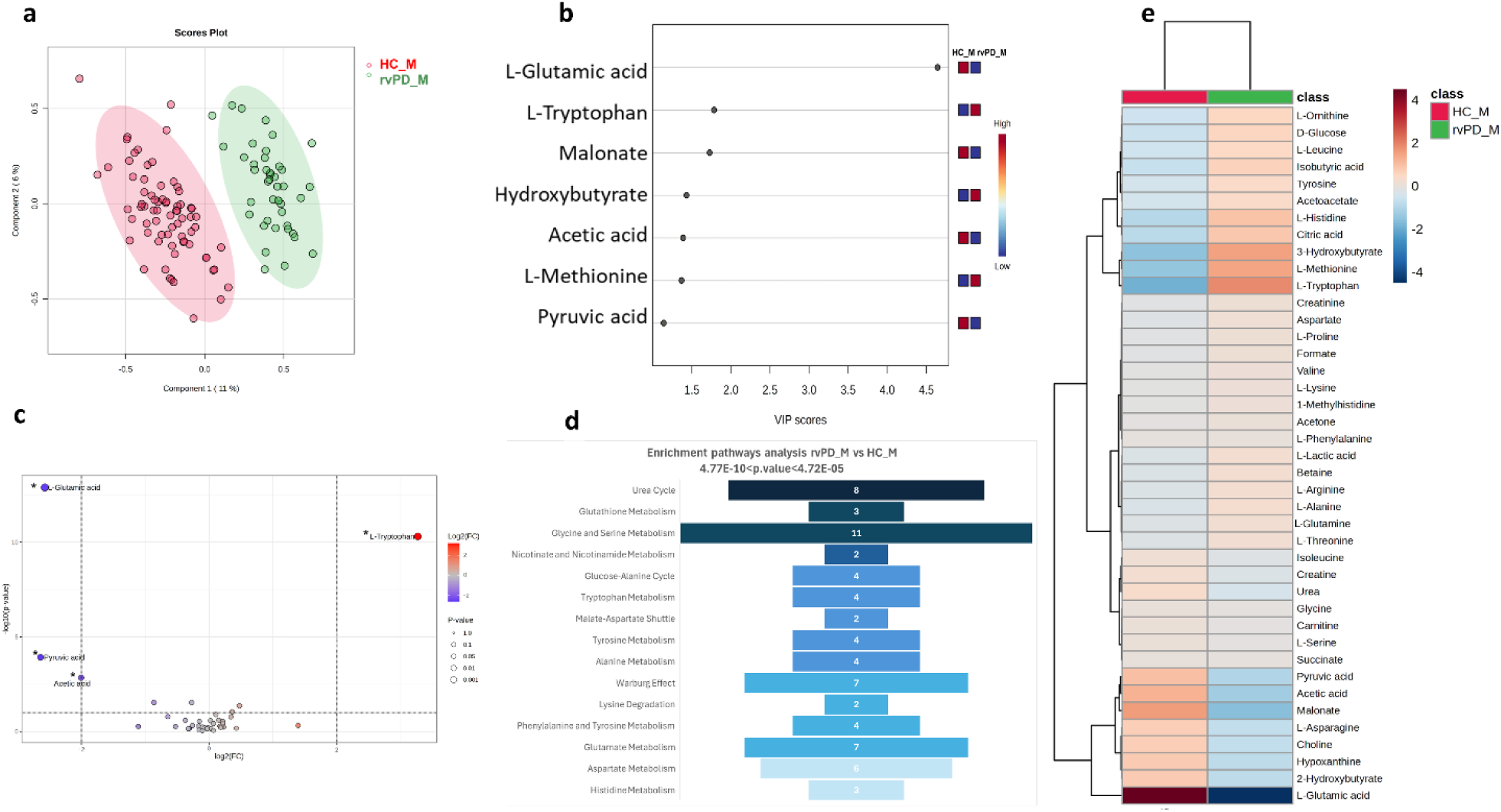
Male gender influences serum metabolome profiles in PD patients with rare genetic variants compared with healthy controls. **a**, The Partial Least Squares Discriminant Analysis (PLS-DA) score plots of serum metabolite levels from male rvPD patients (44 patients, red; 77 healthy controls, green). Variance explained: PC1 = 11%, PC2 = 6%. Models were validated by 10-fold CV (accuracy = 0.96, 0.99; Q² = 0.73, 0.79). **b,** Variable Importance Projection (VIP) plots reporting the metabolites responsible for cluster separations. Only metabolites with VIP > 1 were considered significant. **c**, Robust volcano plots showing the upregulated and downregulated metabolites in the male rvPD serum, represented in red and blue, respectively. The fold-change value was set to 2, while the p-value threshold was set to <0.05. Metabolites significant for both fold-change and p-value from the t-test are labelled with an asterisk. **d**, Pathway enrichment analysis was performed on ¹H-NMR data related to serum metabolomics results of male rvPD patients compared to their sex-matched healthy controls (HC). The bars represent the hits, i.e., the number of metabolites detected in the spectra and involved in the pathways. Pathways were considered statistically significant with hits > 1, p-value < 0.05, and p-values adjusted using the Holm–Bonferroni test (Holm p) and the False Discovery Rate (FDR) < 1. Darker colours represent lower and, therefore, more significant p-values. The range of significance is shown at the top. The exploration of pathways was carried out using the Small Molecule Pathway Database (SMPDB), selecting *Homo sapiens* as the organism. **e,** Heatmaps of the modified metabolites relative to rvPD males. The colour of each section corresponds to the concentration value of each metabolite, calculated from a normalised concentration matrix (red, upregulated; blue, downregulated).

**Figure 3.**
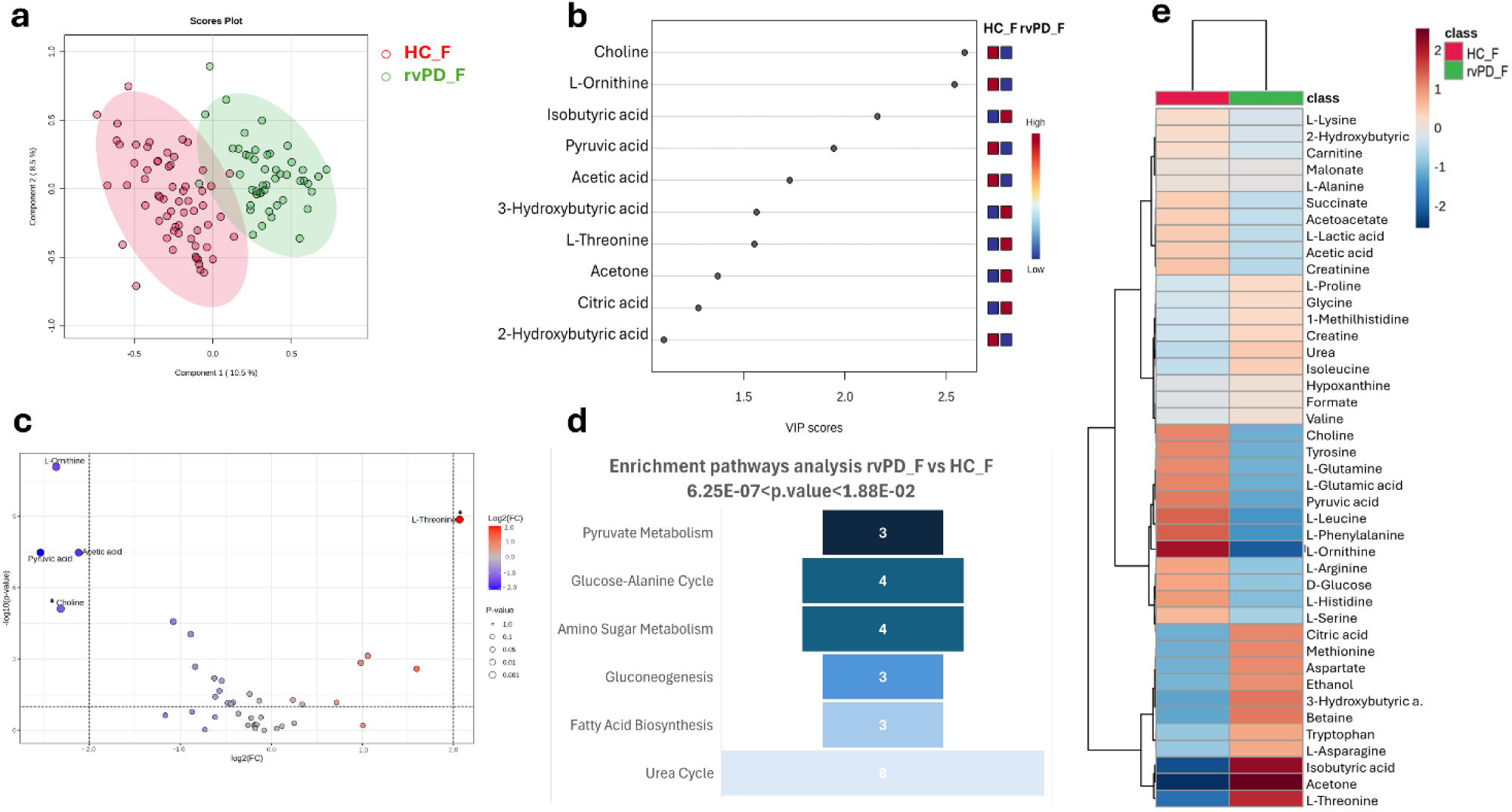
Female gender influences serum metabolome profiles in PD patients with rare genetic variants compared with healthy controls. **a**, The Partial Least Squares Discriminant Analysis (PLS-DA) score plots of serum metabolite levels from female rvPD patients (47 patients, red; 60 healthy controls, green). Variance explained: PC1 = 10.5%, PC2 = 8.5%. Models were validated by 10-fold CV (accuracy = 0.93, 0.95; Q² = 0.64, 0.70). **b,** Variable Importance Projection (VIP) plots reporting the metabolites responsible for cluster separations. Only metabolites with VIP > 1 were considered significant. **c**, Robust volcano plots showing the upregulated and downregulated metabolites in the female rvPD serum, represented in red and blue, respectively. The fold-change value was set to 2, while the p-value threshold was set to <0.05. Metabolites significant for both fold-change and p-value from the t-test are labelled with an asterisk. **d**, Pathway enrichment analysis was performed on ¹H-NMR data related to serum metabolomics results of female rvPD patients compared to their sex-matched healthy controls (HC). The bars represent the hits, i.e., the number of metabolites detected in the spectra and involved in the pathways. Pathways were considered statistically significant with hits > 1, p-value < 0.05, and p-values adjusted using the Holm–Bonferroni test (Holm p) and the False Discovery Rate (FDR) < 1. Darker colours represent lower and, therefore, more significant p-values. The range of significance is shown at the top. The exploration of pathways was carried out using the Small Molecule Pathway Database (SMPDB), selecting *Homo sapiens* as the organism. **e,** Heatmaps of the modified metabolites relative to rvPD females. The colour of each section corresponds to the concentration value of each metabolite, calculated from a normalised concentration matrix (red, upregulated; blue, downregulated).

The sex-stratified multivariate analysis revealed that amino acids had a primary influence in differentiating the serum metabolomic profiles between cases and controls in males. Specifically, VIP analysis indicated the significant discriminative capabilities of L-glutamic acid (VIP: 4.64), L-tryptophan (VIP: 1.78), and L-methionine (VIP: 1.39) in separating the blood metabolomes between patients and controls (**Fig. 2b**).

Besides amino acids, other metabolites contributing to the separation of the serum metabolomic profiles of rvPD males from HCs included malonate (VIP:1.72); 3-hydroxybutyrate (VIP: 1.43), acetic acid (VIP: 1.39) and pyruvic acid (VIP: 1.14) (**Fig. 2b**).

In female patients, VIP analysis identified choline (VIP: 2.59) as the most distinguishing metabolite compared to controls **(Fig. 3b)**. Moreover, L-ornithine (VIP: 2.54) and L-threonine (VIP: 1.55) also differentiated the serum metabolomic profiles between women with rvPD and HCs **(Fig. 3b)**. Additional discriminating molecules between cases and controls included isobutyric acid (VIP: 2.16), pyruvic acid (VIP: 1.94), acetic acid (VIP: 1.72), 3-hydroxybutyrate (VIP: 1.56), acetone (VIP:1.37), citric acid (VIP: 1.27), and 2-hydroxybutyrate (VIP: 1.10) **(Fig. 3b)**.

Notably, in the male rvPD cohort, the robust volcano plot revealed significantly lower concentrations of L-glutamic acid, pyruvic acid, and acetic acid compared to HCs (**Fig. 2c, Table S5**). In contrast, L-tryptophan concentrations were higher in these patients than in healthy individuals (**Fig. 2c, Table S5**).

A significant decrease of pyruvic acid and acetic acid was observed in the serum of female rvPD patients compared to sex-matched HCs (**Fig. 3c, Table S5**). Additionally, the robust volcano plot indicated a female-specific downregulation of choline and L-ornithine levels, as well as an upregulation of L-threonine compared to sex-matched HCs (**Fig. 3c, Table S5**).

Consistent with metabolomic results, biomarker analysis utilizing the Area Under the Curve (AUC) demonstrated that glutamate (AUC = 1.00) and tryptophan (AUC = 0.96) effectively differentiate male rvPD patients from HCs (**Fig. S2**). On the other hand, threonine levels (AUC = 0.90) demonstrated potential as a sex-specific biomarker for rvPD in females (**Fig. S2**).

Afterward, we conducted an enrichment pathway analysis to identify the main cellular pathways involved in the dysregulation of the serum metabolomic profile in individuals with rvPD, comparing male and female patients with sex-matched HCs. Remarkably, in males, data analysis revealed fifteen dysregulated biochemical pathways in cases compared to controls (**Fig. 2d, Table S6**). These biochemical processes included the metabolism of glycine-serine, tryptophan, tyrosine, phenylalanine, alanine, glutamate, aspartate, and histidine, as well as the degradation of lysine. Moreover, patients harbouring rare variants also reported altered urea cycle, glucose-alanine cycle, malate-aspartate shuttle, Warburg effect, nicotinate and nicotinamide metabolism, compared to HCs (**Fig. 2d, Table S6**). Of note, a significant alteration in glutathione biosynthesis was observed among the most prominent biochemical pathways affected in male patients compared to controls (**Fig. 2d**, **Table S6**).

In female patients, enrichment pathway analysis showed only six dysregulated pathways compared to HCs: pyruvate metabolism, glucose-alanine cycle, amino sugar metabolism, gluconeogenesis, fatty acid biosynthesis, and urea cycle (**Fig. 3d, Table S6**).

The heatmap analysis further confirmed the metabolites’ variation identified in the serum of female and male rvPD patients compared to sex-matched controls (**Fig. 2e, 3e).**

Consistent with previous observations ^26,47,48,60,61^, the current untargeted NMR findings in rvPD substantiate a remarkable influence of sex differences in shaping serum metabolite variations compared to matched HCs.

### ^1^H-NMR analysis shows differences in the serum metabolome profiles between male and female patients with rvPD

To further investigate the impact of sex differences on serum metabolomic profiles in patients with rare variants, we analyse the metabolomic features in independent cohorts of HCs (male: 77, female: 60) and rvPD (male: 44, female: 47), stratified by sex.

The supervised PLS-DA analysis demonstrated overlap in the serum metabolomes between male and female healthy subjects, as evidenced by PC1’s Q2 value of 0.003 (**Figure 4a**), indicating the absence of clustering and the homogeneity of the serum metabolome across HCs of both sexes.

**Figure 4.**
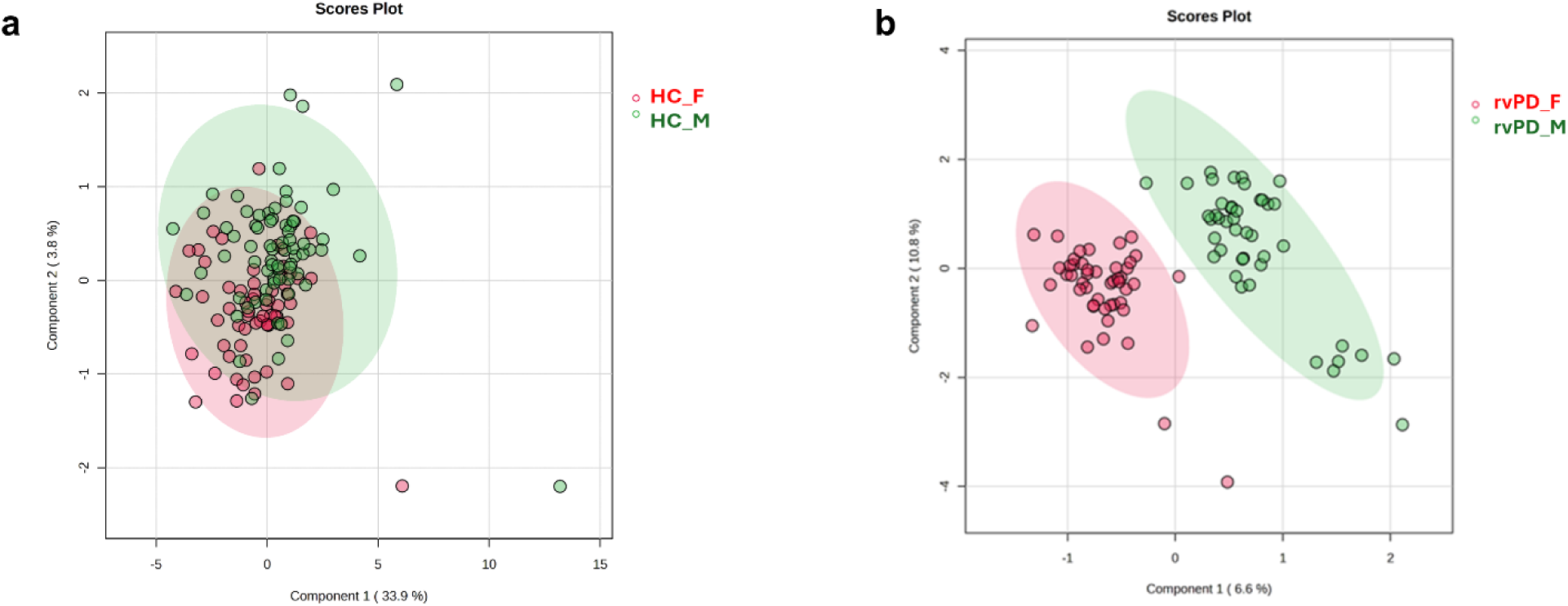
¹H-NMR metabolomics analysis reveals disparities in the serum metabolome profiles between male and female patients diagnosed with rvPD. Partial Least Squares Discriminant Analysis (PLS-DA) score plots are shown in Cartesian space, with the x- and y-axes representing the percentage of variance explained by the first two principal components (**a**: PC1 = 33.9%, PC2 = 3.8%; **b**: PC1 = 6.6%, PC2 = 10.8%).The supervised models were constructed using serum metabolite concentrations from 137 healthy controls (**panel a**: male = 77; green cluster, female = 60; red cluster) and 91 patients carrying rare genetic variants (**panel b**: male = 44, green cluster; female = 47, red cluster). Model validation was performed using a 10-fold cross-validation (CV) procedure, yielding the following performance metrics: (**a**) accuracy = 0.59 and 0.57 for PC1 and PC2, respectively, with Q² values of 0.003 and –0.19; (**c**) accuracy = 0.92 and 0.98, with Q² values of 0.47 and 0.78.

On the other hand, the supervised model applied to the serum metabolome of rvPD patients revealed a significant sex-dependent separation of metabolomic profiles, with PC1 exhibiting a Q2 value of 0.47 (**Figure 4b**). Noteworthy, the analysis of VIP scores and volcano plots revealed that the differences in blood metabolome profiles between male and female rvPD patients are mainly influenced by higher levels of alanine in the female patient group (**Fig S3a,b**). Additionally, compared to male patients, the female group also exhibited increased levels of asparagine and decreased levels of tyrosine (**Fig S3a,c**). The sex-specific metabolic abnormalities observed in male and female patients with rvPD are further validated by pathway-enrichment analysis. In fact, this NMR data analysis reveals variations in the metabolism of selenoamino acids and tryptophan, the biosynthesis of glutathione, and the dysregulation of bioenergetic processes such as the glucose-alanine cycle between the sexes. (**Fig S3d, Table S7**). Importantly, these sex-specific metabolic alterations emerge despite male and female rvPD patients exhibiting comparable clinical and demographic characteristics (**Table S8**), underscoring that the observed biochemical differences are unlikely to be driven by clinical heterogeneity. Importantly, the current data further confirm a key role of sex differences in shaping serum metabolome features in rvPD patients.

### Comparable blood metabolomic profiles across rare variant-, idiopathic-, and genetic-PD patients

Next, we investigated whether the distinct genetic status of PD patients, across both sexes, was associated with specific biochemical variations in their serum metabolome profiles.

Noteworthy, the serum metabolomic profile of rvPD patients in both the entire cohort and sex-stratified groups does not differ from those of gPD and iPD patients **(Fig 5a-f)**.

**Figure 5.**
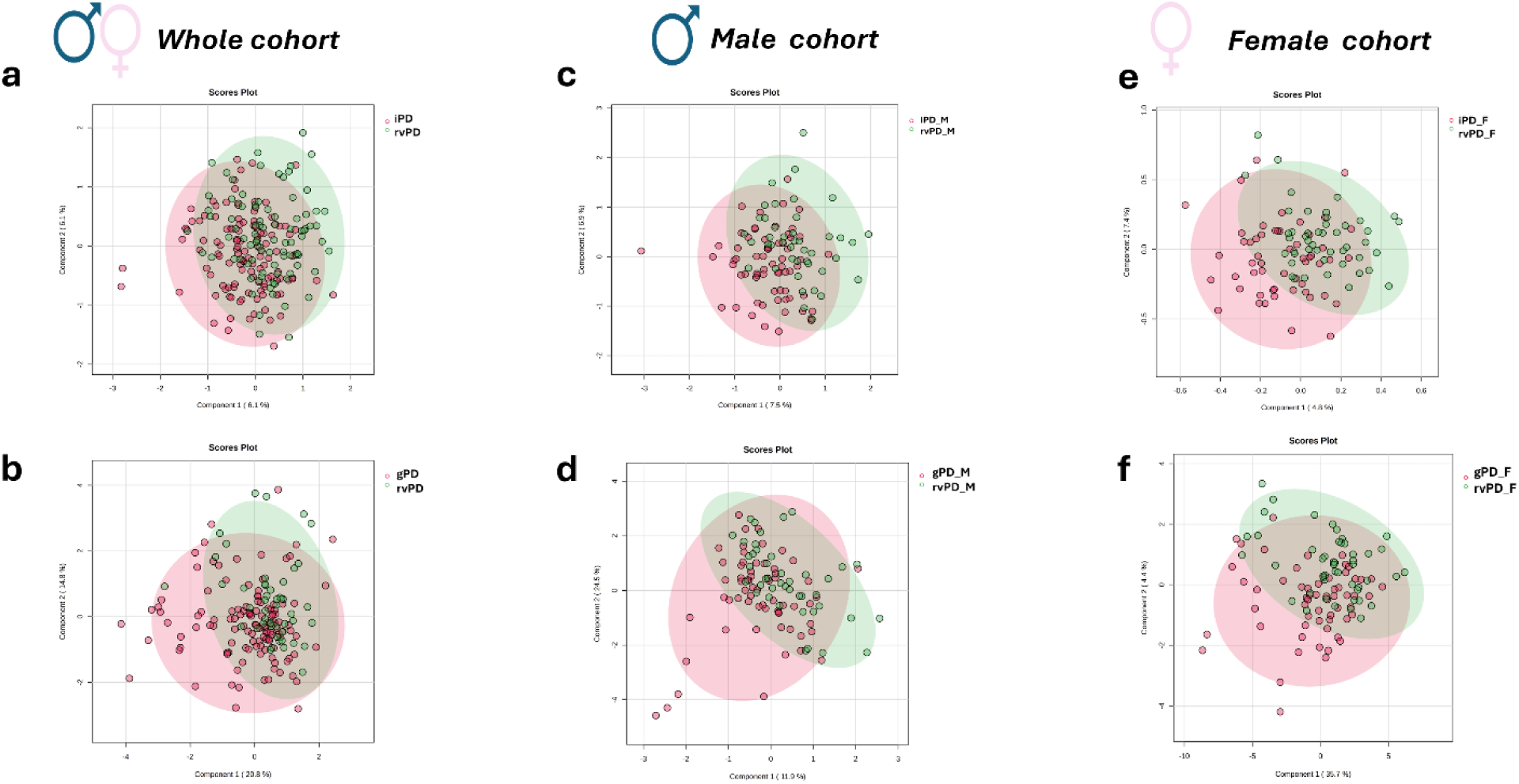
Similar blood metabolomic profiles are observed across rare-variant, idiopathic, and genetic PD patients. **a-f** Partial Least Squares Discriminant Analysis (PLS-DA) score plots are shown in Cartesian space, with the x- and y-axes representing the percentage of variance explained by the first two principal components (iPD vs rvPD: **a**: PC1 = 6.1%, PC2 = 6.1%; **c**: PC1 = 7.5%, PC2 = 6.9%; **e**: PC1 = 4.8%, PC2 = 7.4%; gPD vs rvPD **b**:PC1 = 20.8%, PC2 = 14.8%; **d**:PC1 = 11.9%, PC2 = 24.5%; **f**:PC1 = 35.7%, PC2 = 4.4%). The supervised models were constructed using serum metabolite concentrations from 121 patients with idiopathic Parkinson’s disease (iPD; red clusters; male = 65, female = 56), 124 patients with genetic Parkinson’s disease (gPD; red clusters; male= 64, female =60) and 91 patients carrying rare genetic variants (rvPD; green clusters; male = 44, female = 47). Model validation was performed using a 10-fold cross-validation (CV) procedure, yielding the following performance metrics: (**a**) accuracy = 0.53 and 0.54 for PC1 and PC2, respectively, with Q² values of –0.06 and –0.13; (**c**) accuracy = 0.54 and 0.51, with Q² values of – 0.14 and –0.24; and (**e**) accuracy = 0.46 and 0.48, with Q² values of –0.27 and –0.21; (**b**) accuracy = 0.64 and 0.67 for PC1 and PC2, respectively, with Q² values of –0.02 and –0.10; (**d**) accuracy = 0.49 and 0.53 for PC1 and PC2, respectively, with Q² values of –0.14 for both component; (**f**) accuracy = 0.57 and 0.49 for PC1 and PC2, respectively, with Q² values of –0.03 and –0.18.

Our multivariate NMR analysis showed that PD patients with idiopathic or genetic conditions, as well as those with rare genetic variants - characterized by comparable demographic and clinical features (**Table 1, Table S2**) have similar serum metabolome profiles, regardless of sex.

Our NMR-based metabolomic findings indicate a common systemic metabolic dysregulation across various PD subtypes, regardless of their distinct genetic backgrounds.

### HPLC analysis displays reductions of L-glutamate content in the serum of male patients carrying rare variants compared to matched healthy controls

Based on the notable alteration in the metabolism of glutamate, aspartate, glycine and serine detected by NMR experiments, here we quantified the D-serine (D-Ser), L-serine (L-Ser), glycine (Gly), L-glutamate (L-Glu), L-glutamine (L-Gln), L-aspartate (L-Asp), and L-asparagine (L-Asn) concentrations in rvPD patients and HCs by HPLC analysis (**Fig. 6a**). In addition, we calculated the D-Ser/total Ser and L-Gln/L-Glu ratios as indices of D-Ser and L-Glu biosynthesis from their respective precursors, L-Ser and L-Gln.

**Figure 6.**
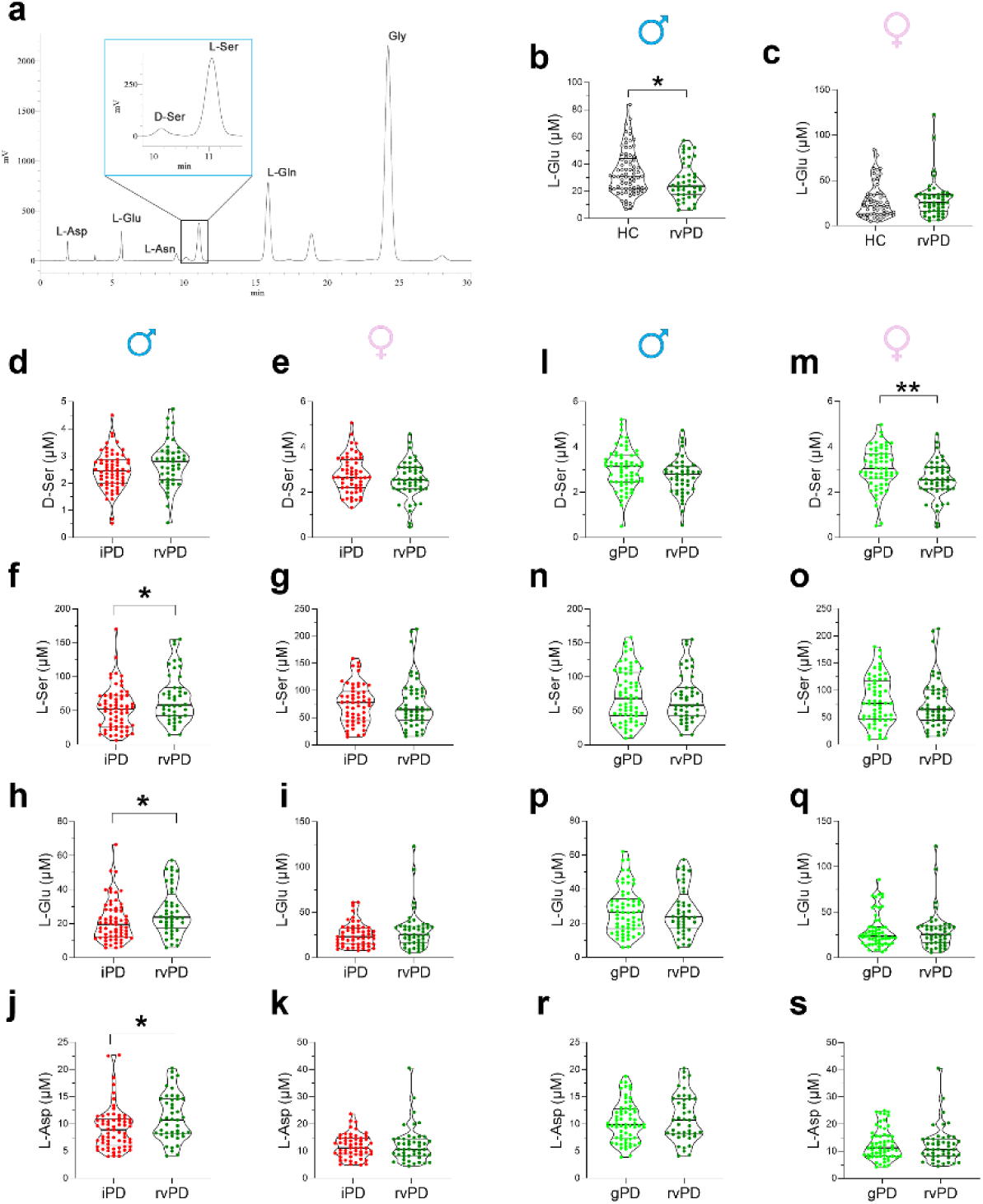
D-serine, L-serine, L-glutamate and L-aspartate levels in the serum of male and female Parkinson’s disease (PD) patients with different PD subtypes. **a,** Representative HPLC chromatogram displaying D-serine (D-Ser), L-serine (L-Ser), L-glutamate (L-Glu), L-glutamine (L-Gln), L-aspartate (L-Asp), L-asparagine (L-Asn), and glycine (Gly) peaks obtained from a serum sample of a PD patient. (**b, c**) Serum levels of L-glutamate (L-Glu) in (**b**) male (n = 44) and (**c**) female (n = 47) rvPD patients, compared to sex-matched healthy controls (HC) (males, n = 77; females, n = 60). (**d-k**) Serum concentrations of (**d, e**) D-Ser, (**f, g**) L-Ser, (**h, i**) L-Glu and (**j, k**) L-Asp compared between patients with idiopathic PD (iPD; males, n = 65; females, n = 56) and in those carrying rare genetic variations (rvPD; males, n = 44; females, n = 47) in male and female subgroups. (**l-s**) Serum concentrations of (**l, m**) D-Ser, (**n, o**) L-Ser, (**p, q**) L-Glu and (**r, s**) L-Asp compared between PD patients carrying pathogenic mutations (gPD; males, n = 64; females, n = 60) and in those carrying rare genetic variations (rvPD; males, n = 44; females, n = 47) in male and female subgroups. In each sample, free amino acids were detected in a single run. Dots represent the single subjects’ values, while lines illustrate the median with interquartile range. *p <0.05, **p < 0.01 (Mann–Whitney U test).

Without sex stratification, no significant changes in serum D- and L-amino acid levels were found between rvPD patients and HCs (**Table S9**). In line with NMR data (**Fig. 2b,c**), sex-stratified analyses revealed a significant reduction of L-Glu content in male rvPD but not females, compared with their respective sex-matched (**Fig. 6b,c**; **Table 2**). This effect remained significant after adjustment for age and LEDD in ANCOVA on the original, non-transformed variable (**Table 2**).

Although male rvPD patients exhibited a notable trend toward reduced levels of L-Ser, Gly, and L-Asn relative to controls, these differences did not reach statistical significance (**Table 2**). All other amino acid concentrations were comparable between groups (**Table 2**). Unlike males, no significant differences in D- and L-amino acid levels were observed between female rvPD patients and sex-matched HCs (**Table 2**).

After, we investigated whether the genetic status of male and female patients with distinct PD subtypes differentially affects the circulating levels of the D- and L-amino acids, as previously evaluated between cases and controls.

HPLC analysis reveals a significant reduction in L-Ser, L-Glu, L-Asp and L-Asn levels in the blood of male iPD patients compared with sex-related rvPD. (**Fig. 6f,h,j**; **Table 3**). These alterations, except for L-Asn, were further confirmed by age-, LEDD- and disease duration-adjusted ANCOVA analyses on natural log-transformed data (**Table 3**). In contrast, the levels of all other amino acids did not differ significantly between iPD and rvPD subgroups (**Table 3**). Unlike males, in females no significant differences in amino acid content were observed between iPD and rvPD patients (**Fig. 6e,g,i,k**; **Table 3**).

**Table 3.** Concentrations of serum D- and L- amino acids content compared between idiopathic PD and PD patients carrying rare variants in male and female subgroups.

| Sex | Amino acids | iPD |  |  |  | rvPD |  |  |  | Mann-Whitney | ANCOVA<sup>a</sup> |  |
| --- | --- | --- | --- | --- | --- | --- | --- | --- | --- | --- | --- | --- |
|  |  | N | Median | IQR |  | N | Median | IQR |  | *p* value | F<sub>(1;104)</sub> | *p* value |
| Male | L-Asp (μM) | 65 | 8.9 | 6.3 | 10.8 | 44 | 10.7 | 8.2 | 14.6 | **0.010** | 4.88 | **0.029** |
|  | L-Glu (μM) | 65 | 19.5 | 12.5 | 28.1 | 44 | 23.7 | 17.5 | 35.6 | **0.022** | 4.50 | **0.036** |
|  | L-Asn (μM) | 65 | 14.8 | 8.5 | 20.8 | 44 | 18.3 | 10.8 | 27.4 | **0.047** | 3.52 | 0.064 |
|  | D-Ser (μM) | 65 | 2.5 | 2.0 | 2.9 | 44 | 2.8 | 2.1 | 3.1 | 0.068 |  |  |
|  | L-Ser (μM) | 65 | 52.6 | 26.8 | 71.7 | 44 | 58.3 | 42.6 | 83.9 | **0.027** | 4.54 | **0.035** |
|  | D-Ser/total Ser (%) | 65 | 4.2 | 3.4 | 7.5 | 44 | 3.9 | 2.6 | 5.6 | 0.093 |  |  |
|  | L-Gln (μM) | 65 | 132.8 | 90.0 | 181.5 | 44 | 157.7 | 107.2 | 194.8 | 0.068 |  |  |
|  | Gly (μM) | 65 | 111.6 | 78.5 | 149.1 | 44 | 123.9 | 94.1 | 149.9 | 0.138 |  |  |
|  | L-Gln/L-Glu | 65 | 6.5 | 5.1 | 9.0 | 44 | 6.5 | 5.1 | 8 | 0.693 |  |  |
| Female | L-Asp (μM) | 56 | 11.2 | 8.9 | 14.7 | 47 | 10.7 | 8.3 | 14.6 | 0.667 |  |  |
|  | L-Glu (μM) | 56 | 23.0 | 14.8 | 32.5 | 47 | 25.5 | 16.2 | 34.6 | 0.391 |  |  |
|  | L-Asn (μM) | 56 | 20.0 | 12.1 | 27.3 | 47 | 22 | 12.4 | 28.3 | 0.689 |  |  |
|  | D-Ser (μM) | 56 | 2.7 | 2.2 | 3.4 | 47 | 2.5 | 2.1 | 3.1 | 0.232 |  |  |
|  | L-Ser (μM) | 56 | 77.6 | 45.7 | 97.8 | 47 | 65 | 44.9 | 100.1 | 0.617 |  |  |
|  | D-Ser/total Ser (%) | 56 | 3.5 | 3.0 | 4.4 | 47 | 3.6 | 2.6 | 4.9 | 0.812 |  |  |
|  | L-Gln (μM) | 56 | 188.5 | 129.3 | 231.3 | 47 | 176.3 | 119.4 | 233.4 | 0.791 |  |  |
|  | Gly (μM) | 56 | 173.8 | 110.6 | 216.7 | 47 | 158.8 | 115.8 | 214.2 | 0.905 |  |  |
|  | L-Gln/L-Glu | 56 | 7.4 | 5.8 | 9.6 | 47 | 6.7 | 5.5 | 8.7 | 0.223 |  |  |

Abbreviations: iPD, idiopathic Parkinson's disease; rvPD, Parkinson's disease with rare variants; N, number of subjects; IQR, interquartile range.
<sup>a</sup> adjusted for age, LEDD and disease duration

We next compared the serum levels of D- and L-amino acids between rvPD and gPD patients across both sexes. Overall, non-parametric Mann-Whitney analysis failed to detect significant differences in amino acid levels between rvPD and gPD patients in both male and female subgroups (**Fig. 6l-s**, **Table 4**). However, a selective reduction in D-Ser levels emerged in female patients with rvPD compared to those with gPD (**Fig. 6l,m**, **Table 4**). This finding was confirmed through adjusted ANCOVA analyses considering age, LEDD and disease duration as covariates (**Table 4)**. Overall, the current HPLC findings further confirm a remarkable influence of sex differences and genetic background on circulating D- and L- amino acid profiles in PD patients.

**Table 4.**
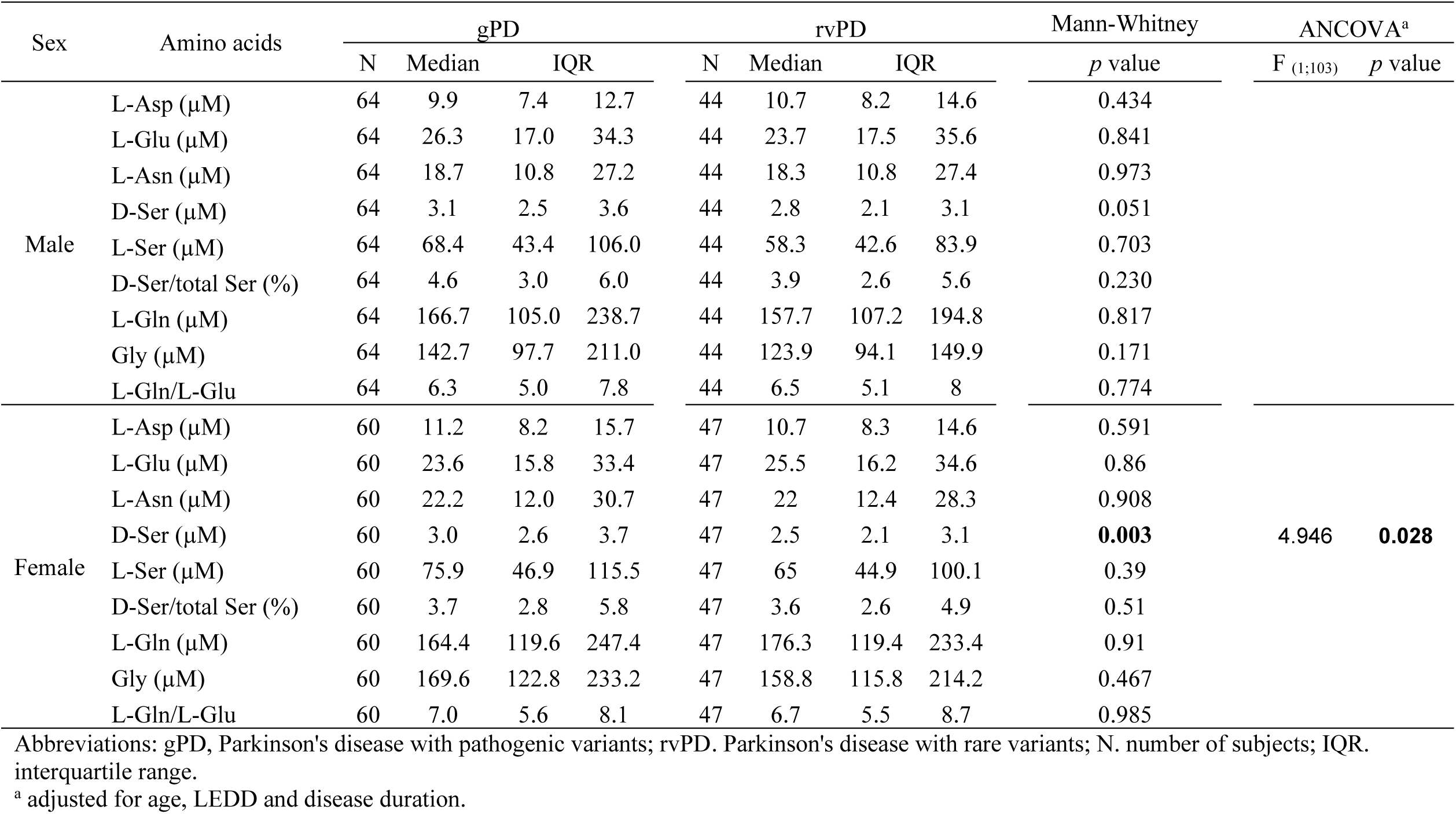
Concentrations of serum D- and L- amino acids content compared between genetic PD and PD patients carrying rare variants in male and female subgroups.

### Association of amino acid concentrations with age, age at onset, duration of disease, and antiparkinsonian treatment in PD patients harbouring rare genetic variants

Next, we examined potential relationships between NMDAR-related amino acids, their precursors, and demographic or clinical variables, including age, age at onset, disease duration, and antiparkinsonian treatment expressed as LEDD, in male and female rvPD patients (**Table 5; Table S10**). Overall, Spearman’s correlation analysis revealed no significant associations between serum D- and L-amino acid levels and age in either sex, except for a significant positive correlation between the L-Gln/L-Glu ratio and age in female patients (**Table 5**). In addition, a negative association between L-Glu levels and age at PD onset was observed in female rvPD patients, whereas no significant correlations were found for the other amino acids (**Table S10)**. Furthermore, no significant associations with disease duration or LEDD were observed in rvPD patients after adjusted partial correlation analysis, irrespective of sex. (**Table S10)**.

**Table 5.** Correlation of serum D- and L- amino acids levels with the age of male and female PD patients carrying rare variants.

| Sex | Parameter | L-Asp (μM) |  | L-Glu (μM) |  | L-Asn (μM) |  | D-Ser (μM) |  | L-Ser (μM) |  | D-Ser/total Ser (%) |  | L-Gln (μM) |  | Gly (μM) |  | L-Gln/L-Glu |  |
| --- | --- | --- | --- | --- | --- | --- | --- | --- | --- | --- | --- | --- | --- | --- | --- | --- | --- | --- | --- |
|  |  | r | *p* value | r | *p* value | r | *p* value | r | *p* value | r | *p* value | r | *p* value | r | *p* value | r | *p* value | r | *p* value |
| Male (N=44) | Age (years) | 0.148 | 0.339 | 0.226 | 0.140 | 0.141 | 0.360 | 0.243 | 0.111 | 0.135 | 0.384 | -0.036 | 0.814 | 0.161 | 0.295 | 0.128 | 0.408 | -0.090 | 0.560 |
| Female (N=47) | Age (years) | -0.217 | 0.143 | -0.252 | 0.087 | -0.127 | 0.395 | -0.075 | 0.615 | -0.096 | 0.522 | 0.061 | 0.683 | 0.043 | 0.775 | 0.072 | 0.631 | 0.421 | **0.003** |

Abbreviations: N, number of subjects. Significant p values from Spearman's correlation analyses are shown in bold

### Sex-dependent associations between serum amino acid levels and motor symptoms score of PD patients carrying rare variants

Here, we investigated the potential associations between serum levels of NMDAR-related amino acids and their direct precursors with motor symptoms, evaluated by MDS-UPDRS III scores in male and female rvPD patients. In males, Spearman ‘correlations analysis revealed positive associations between serum levels of L-Glu, L-Ser, L-Asp and Gly and MDS-UPDRS III scores (**Fig. 7a, c, g, i**; **Table 6**). In contrast, among female rvPD patients, positive correlations were observed between L-Ser and L-Gln content and MDS-UPDRS III scores (**Fig. 7d, f**; **Table 6**), with no significant correlations observed with the other D- and L-amino acids analyzed (**Fig. 7b, h, j**; **Table 6**). All these associations remained statistically significant after partial correlation analysis adjusted for age, disease duration, and LEDD.

**Figure 7.**
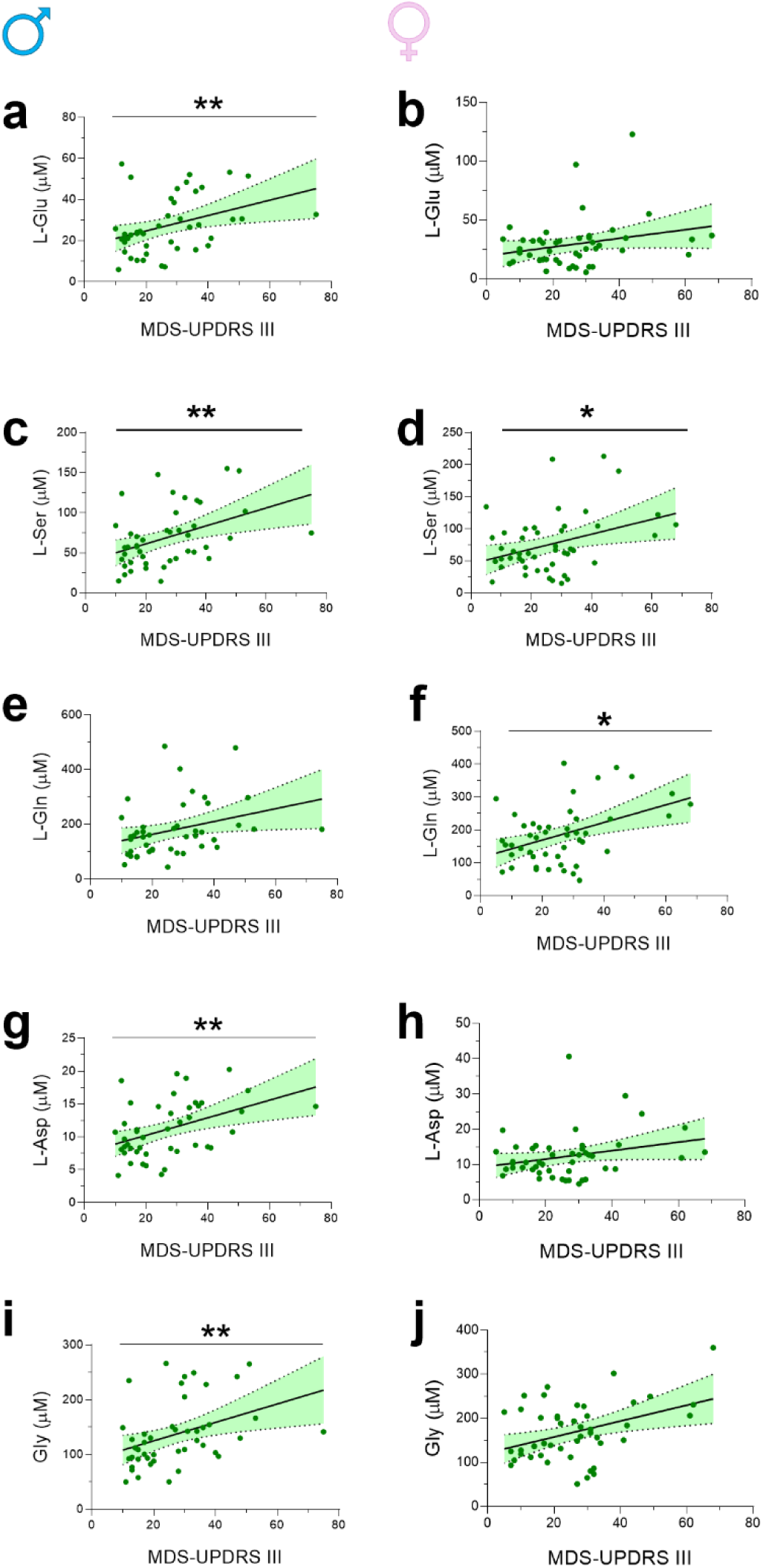
Correlations of serum amino acids content with MDS-UPDRS III scores in PD patients carrying rare genetic variants. Scatterplots showing the correlations between serum (**a, b**) L-glutamate (L-Glu), (**c, d**) L-serine (L-Ser), (**e, f**) L-glutamine (L-Gln), (**g, h**) L-aspartate (L-Asp) and **(i, j**) glycine (Gly) levels and motor symptom severity, as assessed by MDS-UPDRS Part III scores in male (n = 44) and female (n = 47) Parkinson’s disease patients carrying rare genetic variants. Lines and green shadows represent the best fit line and its 95% CI, respectively. *p* <0.05, ** *p* < 0.01, Spearman’s correlation test (confirmed by partial correlation analysis adjusted for age, disease duration, and LEDD).

**Table 6.** Correlation of serum D- and L- amino acids levels with motor symptoms scores of male and female PD patients carrying rare genetic variants.

| Sex | Parameters | L-Asp (μM) |  | L-Glu (μM) |  | L-Asn (μM) |  | D-Ser (μM) |  | L-Ser (μM) |  | D-Ser/total Ser (%) |  | L-Gln (μM) |  | Gly (μM) |  | L-Gln/L-Glu |  |
| --- | --- | --- | --- | --- | --- | --- | --- | --- | --- | --- | --- | --- | --- | --- | --- | --- | --- | --- | --- |
|  |  | r | p value | r | p value | r | p value | r | p value | r | p value | r | p value | r | p value | r | p value | r | p value |
| Male (N=44) | MDS-UPDRS III | 0.426 | **0.004** | 0.408 | **0.006** | 0.431 | 0.003 | 0.298 | 0.049 | 0.43 | **0.004** | -0.203 | 0.187 | 0.402 | 0.007 | 0.467 | **0.001** | -0.046 | 0.765 |
| Female (N=47) | MDS-UPDRS III | 0.201 | 0.176 | 0.242 | 0.101 | 0.298 | 0.042 | 0.152 | 0.308 | 0.29 | **0.048** | -0.173 | 0.245 | 0.35 | **0.016** | 0.248 | 0.093 | 0.074 | 0.623 |

Abbreviations: N, number of subjects; MDS-UPDRS III, Movement Disorders Society Unified Parkinson's Disease Rating Scale, part III.
*p* values shown in bold were confirmed by partial correlation analyses controlling for age, disease duration, and LEDD.

These findings highlight a sex-specific association between serum amino acid levels and PD motor symptoms, with the most significant associations observed in male rvPD patients.

### Genetic variations in *SHMT1*, *SHMT2* and *GCSH* are associated with different PD subtypes and sex among individuals

Recently, we reported that variations in genes related to glycine-serine metabolism (*PHGDH, DAO, SRR,* and *GCSH*) and those encoding NMDAR subunits (*GRIN2A*) act as genetic modifiers for PD in a specific cohort of idiopathic and genetic patients stratified by sex ^45,47^. Building on these findings and considering the glycine-serine metabolism alterations found in male rvPD patients (**Fig. 2d**), we next investigated whether variations in the same genes are associated with PD in patients with rare variants stratified by sex.

A total number of 150 variants with Minor Frequency Allele (MAF) > 0.01 (MAF was referred to our internal database) were selected in the genomic regions from start to end (based on Genome Reference Consortium Human Build 38 (GRCh38) Whole Exome Sequencing, WES, data) of *SRR, DAO, DAOA, SHMT1, SHMT2, PHGDH, AMT, GCSH, GLDC, GRIN1, GRIN2A* and *GRIN2B* genes ^62–67^ including exonic regions, 5’ and 3’ untranslated regions (UTRs), and intronic regions surrounding exon/intron boundaries.

The case-control association test was performed on the same cohort used for NMR and HPLC analyses, including 91 rvPD patients and 137 HCs. The additive genetic model association test was performed with PLINK2 software, correcting for age and 10 Principal Components (PC) to adjust for genetic differences among individuals. Results identified a significant association with PD for the two intronic variants rs2273028 (p = 0.018, Odd Ratio (OR) = 11.7) and rs34095989 (p = 0.02, OR = 2.19) in *SHMT1* and *SHMT2* genes, respectively, by comparing the subgroup of male rvPD subtype with sex-matched HCs (**Table 7**). On the contrary, the two intronic variants rs8177904 (p = 0.01, OR = 4.48) and rs11055581 (p = 0.004, OR = 2.42) in *GCSH* and *GRIN2B* genes were associated with PD in the subgroup of women patients carrying rare variants (**Table 7**). Since the rs11055581 variant in *GRIN2B* has been identified in our previous study as risk factor for PD in a large, unstratified, case-control cohort ^47^, it was excluded from further analyses.

**Table 7.** Association analysis of common variants in *SHMT1*, *SHMT2* and *GCSH* with PD in patients carrying rare variants stratified by sex.

| Sex<br>(Cohort) | Comparison<br>(N) | Gene | chr | Position<br>hg38 | nt<br>change | position | SNP id | test | p-value | OR |
| --- | --- | --- | --- | --- | --- | --- | --- | --- | --- | --- |
| **Male** | rvPD vs HC | SHMT1 | 17 | 18335698 | G>A | Intronic | rs2273028 | ADD | 0.018 | 11.7 |
| (PD-cohort vs HC-cohort) | (45 vs 77) | SHMT2 | 12 | 57233934 | G>A | Intronic | rs34095989 | ADD | 0.02 | 2.19 |
| **Female** | rvPD vs HC | GCSH | 16 | 81087862 | A>G | Intronic | rs8177904 | ADD | 0.01 | 4.48 |
| (PD-cohort vs HC-cohort) | (47 vs 60) | GRIN2B | 12 | 13675725 | C>T | Intronic | rs11055581 | ADD | 0.004 | 2.42 |
| **Male** | rvPD vs HC | SHMT1 | 17 | 18335698 | G>A | Intronic | rs2273028 | ADD | **0.0003\*** | 12.69 |
| (MNI-rvPD vs MNI-HC) | (236 vs 108) | SHMT2 | 12 | 57233934 | G>A | Intronic | rs34095989 | ADD | 0.05 | 1.51 |
| **Female** | rvPD vs HC | GCSH | 16 | 81087862 | A>G | Intronic | rs8177904 | ADD | 0.02 | 3.96 |
| (MNI-rvPD vs MNI-HC) | (135 vs 174) |  |  |  |  |  |  |  |  |  |

PD, Parkinson's disease; MNI-HC, Healthy Controls from MNI cohort genetic biobank; PD-cohort and HC-cohort, PD patients and healthy controls used in Nuclear Magnetic Resonance/High-performance liquid chromatography analyses; rvPD, PD patients carrying rare variants in PD genes; p, p-value calculated with logistic regression model with Plink software; ADD, additive genetic model; OR, Odds Ratio; \*Significance after Bonferroni correction $p=3.3E-04$ (150 variants). Genes tested in the association analysis: *SRR*, *DAO*, *DAOA*, *SHMT1*, *SHMT2*, *GLDC*, *AMT*, *GCSH*, *PHGDH*, *GRIN1*, *GRIN2A*, *GRIN2B*

Considering the limited number of samples used for NMR/HPLC experiments, we validated these genetic associations in a large independent cohort stratified by sex and genetic background that was selected from our entire Italian cohort (Mediterranean Neurological Institute (MNI)). Specifically, 371 sex-stratified rvPD patients (MNI-rvPD, 236 males and 135 females) were compared to 282 sex-matched healthy subjects (MNI-HC, 108 males and 174 females) (see Methods for details). We confirmed the association of the two variants rs2273028 in *SHMT1* (p = 0.0003, OR = 12.6) and rs34095989 (p = 0.05, OR = 1.51) in *SHMT2*, in male rvPD patients, as well as of the variant rs8177904 (p = 0.02, OR = 3.96) in *GCSH* in female rvPD patients. Overall, these results support the role of these variants as sex-specific modifiers factors in rvPD patients (**Table 7**). Notably, rs2273028 in *SHMT1* remained significant after Bonferroni correction for multiple testing (corrected threshold p < 3.3E-04 (0.05/150 variants)).

Subsequently, we investigated if the three identified variants rs2273028, rs34095989 and rs8177904 in *SHMT1*, *SHMT2* and *GCSH*, respectively, could act as susceptibility or protective factors for PD without applying sex and genetic stratification. We performed case-control association study in two independent large cohorts of patients by comparing the 804 patients of the entire MNI-PD cohort with the 282 healthy individuals of the MNI-HC cohort and the 4,586 PD patients of the Parkinson’s disease Genetic Consortium (PDGC) with the 43,989 individuals from the general population of the United Kingdom (UK) biobank, without applying sex and genetic stratification. The analysis PDGC versus UK biobank was performed with Fisher exact test because for this study cohort only aggregated genotypes were available (https://pdgenetics.shinyapps.io/VariantBrowser/). None of the identified variants was associated with PD in these large case-control cohorts (**Table S11**), suggesting that *SHMT1*, *SHMT2* and *GCSH* are not susceptibility genes for PD. However, the potential role of these variants as genetic modifying factors for the rvPD subtype stratified by sex should be interpreted with caution due to the small size of the dataset and needs to be validated in a larger study cohort.

### The rs2273028 variant in SHMT1 gene is associated with increased gene expression in basal ganglia of human brain

To better understand the functional role of the variants in *SHMT1*, *SHMT2* and *GCSH* associated with PD in rvPD patients stratified by sex, we surveyed the Genotype-Tissue Expression (GTEx) portal (https://www.gtexportal.org/home/), to explore the impact of the identified variants on gene expression in various human brain areas including: Amygdala, Anterior cingulate cortex BA24, Caudate basal ganglia, Cerebellum, Cortex, Frontal Cortex BA9, Hippocampus, Hypothalamus, Nucleus accumbens basal ganglia, Putamen basal ganglia, Substantia nigra. Data collected in this database are not stratified for sex and PD subtype. Notably, we identified that the risk allele rs2273028-A in *SHMT1*, linked with PD only in male rvPD patients, was significantly associated with increased expression of *SHMT1* transcript in the Amygdala (p = 7.6E-04), Anterior cingulate cortex (B24) (p = 3.1E-07), Nucleus Accumbens (p = 1.2E-05) and Caudate (p = 3.1E-08) (**Fig. 8; Table S12**).

**Figure 8.**
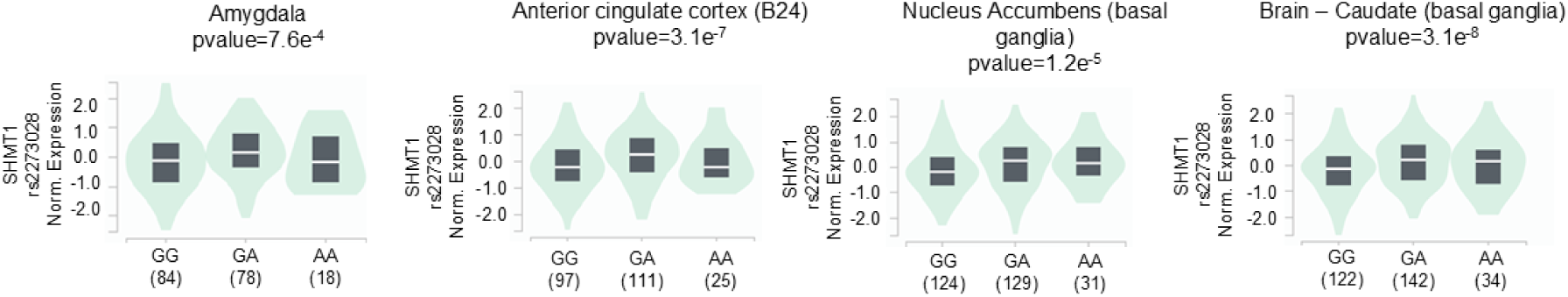
The expression of *SHMT1* transcript in human brain regions is associated with the presence of the risk allele rs2273028-A. Violin plots show significantly increased expression of *SHMT1* transcript associated with the presence of the risk allele rs2273028-A in Amygdala, cortex, Nucleus Accumbens and Caudate. On the x-axis we reported the number of individuals (in brackets) carrying the specific genotype.

## Discussion

Accumulating metabolomic findings have indicated pronounced alterations in amino acids, lipids and sugars, antioxidant molecules, and energy-related metabolite levels in the blood of patients with PD ^25,26,28,30,31,68–74^. However, inconsistencies in the variation and magnitude of these changes — due to heterogeneous analytical platforms, non-standardised protocols, and small cohort sizes — have hindered clear biological interpretation ^73^.

Further reflecting the complexity of this neurodegenerative disorder, recent evidence put forward that both sex and genetic background critically influence serum metabolic profiles in both iPD and gPD patients carrying at least one pathogenic mutation in *LRRK2*, *PARK2*, *PINK1*, *PARK7*, *TMEM175*, or *GBA1* genes, relative to sex-matched HCs ^45,47^.

Beyond these well-characterised pathogenic mutations, emerging evidence supports a polygenic model of PD, in which rare variants of uncertain pathogenicity—affecting Mendelian PD genes as well as risk-associated loci—contribute to the genetic heterogeneity observed in familial and sporadic cases ^15,16,75^. In our cohort, patients carried one or more rare variants in genes involved in mitochondrial metabolism, oxidative stress responses, vesicular trafficking, microtubule dynamics, and lysosome–autophagy pathways, all of which are known to be disrupted in PD pathophysiology ^15,16,76–88^. However, the overall impact of these rare genetic variants on circulating metabolome features remains unexplored, making it unclear whether rvPD patients exhibit specific serum alterations relative to HCs and whether their metabolome profile signatures overlap with or differ from those of individuals with iPD or gPD.

For the first time, our NMR-based multivariate analysis robustly distinguished the serum profiles of rvPD patients from those of HCs. Significantly, pathway enrichment analysis revealed that, in comparison to HCs, the metabolic alterations associated with rvPD encompass a broad range of cellular processes involving amino acids metabolism (such as aspartate, alanine, cysteine, glutamate, glycine, serine, phenylalanine, tyrosine, and tryptophan metabolism), vitamin pathways (including folate and nicotinate-nicotinamide metabolism), and the glutathione–mediated antioxidant defence system. Additional metabolic disturbances were observed in purine turnover, short-chain fatty acid metabolism (propionate), amino-sugar pathways, bioenergetic processes (Warburg effect and the glucose–alanine cycle), and nitrogen metabolism, as indicated by dysregulation of the urea cycle and ammonia-recycling. Thus, the identification of multiple altered cellular pathways in the serum of rvPD patients significantly extends previous NMR-based metabolomic findings in both iPD and gPD patients ^47^, further reinforcing the view that regardless of the genetic status, PD is a complex, multisystemic and heterogeneous disorder that emerges beyond the basal ganglia to involve peripheral organs ^89,90^.

While future studies in preclinical and *in vitro* settings are needed to establish the exact causal relationship between genotype and biochemical phenotype, it is important to remark that the rare variants identified in our cohort of patients converge on a limited number of PD-associated genes. In particular, rare variations in genes involved in mitochondrial metabolism may explain the alterations in energy-related metabolites (e.g., pyruvate and ketone bodies) and pathways such as the Warburg effect and glucose-alanine cycle, reflecting a shift toward inefficient energy production and compensatory metabolic reprogramming. In parallel, genetic alteration in lysosomal function and autophagy may contribute to abnormalities in amino acid and lipid metabolism. Increased oxidative stress -likely resulting from mitochondrial impairment and reduced antioxidant capacity- may further account for perturbations in glutathione metabolism. Overall, these findings suggest that distinct rare genetic variants can converge on shared cellular pathways, leading to common systemic metabolic alterations.

Remarkably, when analyzing the case and control groups by sex, the NMR-based multivariate analysis consistently distinguished patients with rvPD from the control individuals. Interestingly, while both male and female rvPD patients shared similar demographic and clinical characteristics, significant metabolic differences were observed related to sex. The serum metabolic profiles of male and female rvPD patients exhibited distinct divergences, whereas no such differences were noted among healthy controls. Supporting this finding, comparisons of the metabolic profiles between male and female rvPD patients and their corresponding sex-matched healthy controls revealed several sex-specific metabolic alterations. Notably, only two pathways—the urea cycle and the glucose–alanine cycle—were found to be dysregulated in both sexes.

Among the pathways selectively altered in male rvPD patients, glutathione metabolism emerged as one of the most affected. This observation is consistent with the pronounced reduction in L-Glu detected only in males and aligns well with clinical evidence reporting higher oxidative stress in male patients with PD ^91,92^. In contrast, female rvPD patients showed prominent alterations in fatty acid biosynthesis, reflected by decreased acetic acid and increased levels of isobutyric acid, acetone, and 3-hydroxybutyrate. This imbalance in metabolites related to lipid metabolism aligns with abnormalities previously reported in PD ^30,47,93–96^.

Abnormal lipid accumulation has been shown to trigger protein aggregation, ultimately contributing to Lewy bodies formation ^97,98^, and lipid metabolism disturbances can modulate inflammatory and signalling pathways involved in dopaminergic neurodegeneration^99^. Hence, our current findings indicating dysregulated lipid metabolism in female rvPD subjects are consistent with previous evidence of free fatty acid accumulation at both systemic and central levels in PD patients and experimental models ^26,30,47,100^.

Consistent with pronounced sexual dimorphism in metabolism, several of the significantly altered pathways identified in the non-stratified comparison between rvPD patients and HCs appeared to be largely driven by the marked downregulation of serum L-Glu levels in male rvPD patients, as indicated by VIP scores, volcano plot analysis, and ROC analysis.

Although such pronounced downregulation of glutamic acid may inflate the notable number of impacted pathways, it is remarkable that several of the identified pathways have already been implicated in PD pathophysiology ^28,31,59,101–105^, underscoring the biological relevance of our current metabolomic findings.

For instance, dysregulation of the glutamatergic system—particularly involving NMDAR signalling—has been consistently reported in both clinical and preclinical studies ^106,107^. L-Glu is the principal excitatory neurotransmitter in the central nervous system, acting on both metabotropic and ionotropic receptors, including NMDAR, where it is crucial for synaptic plasticity ^107,108^. Furthermore, L-Glu is the metabolic precursor of γ-aminobutyric acid (GABA) and participates in essential cellular functions such as energy metabolism under hypoglycaemic conditions and the urea cycle ^109^. L-Glu also contributes to systemic homeostasis, including participation in the vitamin K cycle ^110^, and regulation of peripheral organ functions, such as the pancreas, kidney, liver, heart, stomach, and immune system ^111^. Additionally, in pancreatic islets, glutamate is required for the regulation of insulin secretion, including incretin-mediated insulin release ^112^, and may act as a mediator of gastrointestinal vago–vagal reflexes ^113,114^. In addition, L-Glu plays a critical role in redox homeostasis by contributing to the synthesis of glutathione from L-cysteine and Gly ^104,115^. Additionally, in line with reduced L-Glu concentration, rvPD patients also exhibited dysregulation in nicotinate (niacin) and nicotinamide pathways compared to controls. Nicotinate and nicotinamide are metabolized into NAD⁺ and NADP⁺, cofactors essential for redox balance, energy production, DNA repair, and signal transduction ^116,117^. Thus, the perturbation of nicotinate–nicotinamide metabolism observed in rvPD patients corroborates prior preclinical and clinical reports obtained in both iPD and gPD patients ^47,118^. Accordingly, clinical trials investigating NAD⁺-enhancing strategies are currently underway ^119^.

Interestingly, the upregulation of serum L-Gln and L-ornithine, together with the downregulation of L-Glu—key metabolites in the urea cycle— aligns with recent evidence suggesting elevated cerebral urea as a potential pathogenic mechanism in PD with dementia (PDD) ^47,120^.

On the other hand, the notable alterations in pyruvic acid metabolism, Warburg effect, glucose–alanine cycle, and amino sugar metabolism observed in rvPD patients are consistent with the view that impaired cellular bioenergetics underlies PD ^121^.

Beyond L-Glu, our findings also highlighted a dramatic disruption of glycine-serine metabolism in rvPD patients, as previously reported in other PD cohorts ^45,47^. Specifically, both Gly and D-Ser contribute to NMDAR signalling by acting as obligatory co-agonists at the GluN1 subunit of NMDAR ^122,123^. In both animal models of PD and in patients with this neurological disease, enhanced GluN1 activation has been shown to ameliorate motor and non-motor deficits ^124–127^. Unlike D-Ser, Gly also acts as a potent inhibitory neurotransmitter through activation of Gly receptors, which are highly expressed in basal ganglia nuclei ^128^.

An important aspect to consider when interpreting the sexual metabolic dimorphism observed in our cohort of cases and controls is that these sex-specific blood alterations emerged despite male and female patients exhibiting comparable clinical and demographic characteristics, including age, disease duration, age at onset, LEDD, and MDS-UPDRS scores. Although we cannot exclude the possibility that additional clinical indices not assessed in the present study may differ between sexes—and could potentially mirror the metabolic alterations reported here—the mechanisms underlying these biochemical differences remain unclear. Nonetheless, several sex-related factors, including hormonal, genetic, epigenetic, and lifestyle influences, have been proposed to modulate PD epidemiology, clinical features, and treatment response ^26,129,130^. Future metabolic studies integrating targeted and untargeted approaches will be essential to clarify how these factors interact with circulating metabolic pathways in male and female PD patients.

Intriguing, our findings indicate that the genetic background exerts a nuanced influence on serum metabolic profiles in patients with rvPD compared to those with iPD or gPD. Hence, despite their divergent genetic status, supervised PLS-DA failed to distinguish the serum metabolome profiles of these PD subgroups, a finding that remained unchanged after sex stratification. Consequently, our metabolomic findings imply that iPD, gPD, and rvPD patients share a core set of biochemical PD-linked alterations, precluding a significant multivariate analysis separation of their blood metabolome profiles. Nonetheless, given the critical implications of this observation, further metabolomic investigations in independent, genetically stratified cohorts—using multiple and complementary analytical approaches—are warranted.

The current HPLC analysis of serum amino acids involved in glutamatergic transmission validated our untargeted NMR-based metabolomic findings on the same study cohort. Specifically, the marked reduction in L-Glu levels observed in male rvPD patients through NMR analysis was corroborated by HPLC measurements, which confirmed significantly lower L-Glu concentrations in patients compared to sex-matched controls. Further supporting a role for sex and genetic background in modulating circulating amino acid balance in PD, our HPLC results indicate that male iPD patients exhibit the most pronounced alterations in L-Glu, L-Ser, and L-Asp levels compared with rvPD patients. In contrast, female patients—regardless of genetic background—did not show substantial alteration in amino acid levels, with the exception of an increase in D-Ser observed in gPD patients relative to rvPD.

While the present NMR and HPLC findings indicate sex- and genotype-specific biochemical serum signatures, it remains unclear whether the observed alterations reflect central pathological processes within basal ganglia circuits or, more likely, peripheral metabolic dysregulation linked to pathological changes in peripheral organs. This issue is particularly relevant for alterations involving amino acids such as L-Glu, D-Ser, and Gly, which are implicated in PD pathophysiology. Because of their key roles in neurotransmission and neuroinflammation, their concentrations are tightly regulated within the brain. Although efflux from the CNS to the periphery can occur—especially under conditions of blood–brain barrier (BBB) dysfunction—the reverse transport is highly restricted for L-Glu and Gly ^131,132^. These considerations suggest that the alterations observed in these relevant molecules are predominantly peripheral in origin and that establishing a direct relationship with central neurochemical changes remains challenging. In this context, our recent HPLC and targeted UPLC–MS analyses of *post-mortem* caudate–putamen (CPu) tissue and cerebrospinal fluid (CSF) from PD patients and HCs revealed a more restricted pattern of amino acid alterations compared with serum ^50,59^. Specifically, we observed increased D- and L-serine, proline, and L-Asp in the CPu of PD cases relative to controls, together with reduced L-Glu and elevated levels of both serine enantiomers in the CSF ^50,59^.

Notably, our results indicate no strong associations between NMDAR-related amino acids, their precursors, and demographic or clinical variables -including age, age at onset, disease duration, and LEDD- in our cohort of male and female rvPD patients. The only significant correlations were observed in female rvPD patients, with a positive association between the L-Gln/L-Glu ratio and age, and a negative association between L-Glu levels and age at PD onset. These findings suggest that age-dependent dysregulated L-Glu levels may modulate both the timing of disease onset and the biochemical progression of PD in female rvPD patients. In contrast to our previous findings in females with iPD ^45^, we did not observe a correlation between D-Ser levels and either subject’s age or age at onset, indicating a potential influence of genetic background on these parameters.

Of note, our results reveal a pronounced sexual dimorphism in the associations between serum amino acid profiles and MDS-UPDRS III scores. In male patients, higher serum concentrations of excitatory NMDAR-related amino acids—L-Glu and L-Asp—as well as the neuromodulators L-Ser and Gly, were positively correlated with greater motor symptom severity. In contrast, in female patients, only L-Ser and L-Gln showed positive associations with MDS-UPDRS III scores. Hence, these findings further support the role of sex and genetic status as critical factors modulating the relationship between amino acid levels and motor symptom manifestations of PD patients.

In line with the sex-specific biochemical alterations observed in rvPD patients, we identified in the same cohort intronic variants in *SHMT1* and *SHMT2* associated with PD in males, and a variant in *GCSH* associated with PD in females. These findings extend our previous observations in iPD and gPD cases, supporting a role for genetic variation in glycine–serine metabolism and NMDAR-related genes as modifiers of PD ^45,47^. The associations were replicated in an independent MNI cohort but were not detected in larger, unstratified datasets. Notably, the strongest signal was observed for rs2273028 in *SHMT1*, a gene encoding a key enzyme in serine–glycine metabolism, with the risk allele associated with increased brain expression across both cortical and subcortical regions, suggesting a potential contribution to metabolic dysregulation. However, given the relatively small cohort size and the lack of consistent evidence in larger populations, these findings should be interpreted with caution and require validation in larger, sex-stratified cohorts.

While the present study reveals a key role for sex differences and rare genetic variants in regulating the blood metabolome in PD, we acknowledge that our study has various limitations. First, the cross-sectional design precludes causal inference regarding the relationship between amino acid dysregulation and disease progression, underscoring the need for longitudinal investigations. Second, the age difference between cases and controls (median 5–7 years) may have contributed to some of the metabolic alterations detected by NMR analysis, as suggested by previous evidence ^133,134^. Third, the absence of a clear mechanistic understanding to explain the sex-related differences in metabolomic effects observed in PD patients. Fourth, all PD patients were receiving antiparkinsonian medication at the time of sampling, which may confound the interpretation of metabolic changes and limit our ability to disentangle disease-related alterations from treatment effects.

In conclusion, this study provides evidence that rare genetic variants in PD are associated with distinct alterations in the serum metabolome compared with HCs. We also show that sex is a major determinant of these metabolic differences, with male and female rvPD patients displaying clearly divergent serum metabolite profiles related to sex-matched HCs. Moreover, our NMR analyses reveal that rvPD patients share several metabolite changes and pathway abnormalities with iPD or gPD patients, supporting the presence of common systemic biochemical alterations across PD subtypes. HPLC analyses of amino acids involved in glutamatergic transmission corroborated the metabolomic findings, confirming a significant reduction in L-Glu in male rvPD patients compared with sex-matched HCs and revealing additional amino acid differences influenced by genetic background and sex. Furthermore, we identified sex-specific correlations between selected amino acids and MDS-UPDRS III motor scores, highlighting sex-dependent links between peripheral metabolism and clinical severity.

Together, these findings highlight the importance of integrating sex and genetic background into clinical and biomarker studies aimed at refining our understanding of PD-related metabolic alterations.

## Methods

### Study participants

#### PD cohort

This is a case-control observational study. 336 independent and unrelated PD patients were selected for this study. These patients were part of the PD biobank of IRCCS Neuromed/IGB-CNR. Only individuals aged ≥40 years were included. All the subjects were of European ancestry and were evaluated by qualified neurologists of the Parkinson Centre of the IRCCS INM Neuromed from June 2015 to December 2017, and from June 2021 to December 2023, with a thorough protocol comprising neurological examination and evaluation of non-motor domains. Information about family history, demographic characteristics, anamnesis, and pharmacological therapy was also collected (the treatment of the PD groups consisted for the most of a combination of L-DOPA and dopamine agonists) ^15,16^. Clinical criteria for diagnosis required the presence of at least two cardinal motor signs: asymmetric resting tremor, bradykinesia and rigidity, as well as a good response to L-DOPA and absence of other atypical features and causes of parkinsonism. Exclusion Criteria for enrolment were: *i*) pre-existing psychiatric conditions; *ii*) presence of neurodegenerative neurological diseases such as multiple sclerosis, lateral sclerosis amyotrophic, Alzheimer’s disease, neuromuscular pathologies, epilepsy; *iii*) diagnosis of dementia; *iv*) depression; *v*) prolonged intake of anxiolytics, antidepressants, antipsychotics, hypnotic drugs, cognitive stimulants.

The Movement Disorder Society revised version of the Unified Parkinson’s Disease Rating Scale Part III (33 items, maximum score 132; hereafter called UPDRS) ^135^ was used to assess clinical motor symptoms. These included language, facial expressions, tremor, rigidity, agility in movements, stability, gait and bradykinesia. Patients were analyzed during the ON period. This cohort includes 121 iPD, 124 gPD, and 91 rvPD patients, carrying at least one rare variant not yet confirmed for their pathogenicity in PD genes. The PD cohort was used both for NMR and HPLC analyses and for genetic association tests.

#### Ethical Compliance

All procedures involving human participants were approved by the Institutional Review Board of the IRCCS Neuromed Italy. The study protocols N°9/2015, N°19/2020, N°4/2023 have been registered in clinicaltrial.gov with the numbers NCT02403765 (Release Date: 04/01/2015), NCT04620980 (Release Date: 11/03/2020), NCT05721911 (Release Date: 30/01/2023). Clinical investigations were conducted according to the principles expressed in the Declaration of Helsinki. Written informed consent was obtained from all participants.

The research was carried out following the recommendations set out in the Global Code of Conduct for Research in Resource-Poor Settings.

#### Reporting Standards

“Reporting adheres to the STROBE guidelines for case-control studies and MSI guidelines for metabolomics data.“

No formal sample size calculation was performed; the sample size was determined based on the availability of well-characterized subjects from the biobank.

#### HC cohort

137 healthy subjects matched for sex with PD cohort were selected for the study. Only individuals aged ≥40 years were included. All the HCs were negative for mutation/variant in PD genes (*see next paragraph “Genetic stratification of the Study Cohort” for the criteria used for patient selection*). The HCs cohort was used for both NMR and HPLC analyses and for genetic association test.

To minimize potential sources of bias, standardized protocols for clinical assessment, sample collection, and processing were applied. Laboratory analyses were performed on anonymized samples. Sex-matched controls were included to reduce confounding effects.

### Genetic stratification of the Study Cohort

In the last years, our genetic studies disclosed an extreme genetic heterogeneity associated with PD and suggested a polygenic model of inheritance both in familiar and sporadic cases ^15,16^. We demonstrated that the co-inheritance of multiple rare variants in a panel of 37 PD genes may predict disease risk in about 26 % of patients, many of whom would have been classified as idiopathic ^16^. Nevertheless, our data also suggest that even the presence of a single rare variant is associated with an increased risk of developing PD ^16^. The 37 selected genes include those reported in the literature as Mendelian PD genes (*PARK7*/*DJ-1, DNAJC13, DNAJC6, EIF4G1, FBXO7, LRRK2, PARK2, PINK1, VPS35* and *SNCA*) and those reported as PD genes/at risk factors (*AIMP2, ANKK1, ANKRD50, CHMP1A, GBA1, GIGYF2, GIPC1, GRK5, HMOX2, HSPA8, HTRA2, IMMT, KIF21B, KIF24, MAN2C1, PACSIN1, RHOT2, SLC25A39, SLC6A3, SNCAIP, SPTBN1, TMEM175, TOMM22, TVP23A, UCHL1, VPS8* and *ZSCAN21*) ^15,30,46,136–143^. This panel of 37 PD genes is not intended to be exhaustive of all known PD genes or risk factors; rather, it includes those genes that showed a significantly higher prevalence of rare variants in cases compared to controls ^15,16^.

Specifically, genes such as *ATP13A2*, *VPS13C*, *RAB32*, and *TMEM230*, although previously associated with PD, were excluded from the panel because they did not meet these criteria of statistical significance in our dataset. Moreover, no pathogenic mutations (as reported in LOVD v.3.0 - Leiden Open Variation Database, https://www.lovd.nl/) were identified in these genes in our cohort, which included idiopathic PD, rare variant PD, and genetic PD cases. We fully acknowledge that stratification of PD patients based on the presence of at least one rare variant in the listed genes represent a genetically heterogeneous subgroup, with many different variants of varying penetrance. Nonetheless, without functional studies, it is not possible to determine the effect of individual variants.

In this study whole exome sequencing (WES) data of PD patients and HCs were analyzed to identify rare (Minor allele frequency (MAF) < 0.01 referred to gnomAD v.4.1.0 database) exonic variants (including non-synonymous, splicing and indels), for which pathogenic role was not yet confirmed, in the panel of the selected PD genes.

We selected 121 PD patients that we named “idiopathic”, in which no mutation/variant was identified in the selected PD genes.

The second group, that we named “rare variant-PD” (rvPD), includes 91 PD-affected subjects carrying of at least one rare variant in one of the 37 PD genes described before.

In particular, in the selected patients belonging to rvPD subtype we identified rare variants in 24 of the 37 PD genes including *AIMP2, DNAJC13, DNAJC6, EIF4G1, FBXO7, GIPC1, HMOX2, HSPA8, HTRA2, IMMT, KIF21B, KIF24, LRRK2, MAN2C1, PARK2, RHOT2, SLC25A39, SLC6A3, SNCAIP, SPTBN1, TVP23A, UCHL1, VPS35, ZSCAN21* (**Table S1**). These genes impact on mitochondrial metabolism and oxidative stress, vesicular trafficking, microtubule dynamics and lysosome-autophagy ^15,16,76–88^.

40 of the 91-rvPD patients were carriers of one rare variant in one of the 24 PD genes and 51 were carriers of two or more variants in these PD genes (**Table S1**).

The third group that we named “genetic-PD” (gPD) includes 124 PD patients of which 120 carrying at least a pathogenic mutation in one of the most frequently mutated PD genes such as *LRRK2* (17 patients carrying the p.G2019S and p.R1441C mutations), *TMEM175* (33 patients), *GBA1* (30 patients), *PARK2*, *PINK1* and *PARK7* (40 patients with at least 1 mutations in PARK2/PINK1/PARK7 genes), and four patients carrying two mutations in the selected genes such as *GBA1/TMEM175*, *GBA1/PARK2*, GBA1/PINK1 and *TMEM175/PARK2*, respectively (**Table S1**). The healthy subjects (HC cohort) included in the study were negative for mutations/variants in the selected panel of PD genes.

### Collection and storage of serum samples

Blood sampling was performed after a 6-h fasting. Whole blood was collected by peripheral venepuncture into clot activator tubes and gently mixed. Each sample was stored upright for 30 min at room temperature to allow blood to clot, then centrifuged at 2000× g for 10 min at room temperature. Serum was aliquoted (0.5 mL) in polypropylene cryotubes and stored at −80 °C before usage. Unique anonymized codes have been assigned to the samples for processing and subsequent analysis, maintaining the confidentiality of personal data.

### NMR sample preparation

Serum samples were prepared in accordance with the NMR metabolomics sample quality preservation guidelines ^144,145^. To prepare NMR samples, 100 μL of phosphate buffer (0.075 M Na_2_HPO_4_·7H_2_O, 4% NaN_3_, in H2O/D20) was mixed with 100 μL of blood serum and subsequently transferred into a 3 mm NMR tube. For calibration and measurement of NMR spectra, we utilised trimethylsilyl propionic acid sodium salt (0.1% TSP in D_2_O) as an internal reference signal. NMR spectra were acquired on a Bruker DRX600 MHz spectrometer (Bruker, Karlsruhe, Germany), using a 5 mm triple-resonance z-gradient TXI Probe, and TOPSPIN 3.2 software (Bruker Biospin, Fällanden, Switzerland) for data acquisition and processing.

### NMR spectra acquisition

Carr-Purcell-Meiboom-Gill (CPMG) experiments employed a spectral width of 7 kHz along with 32 k data points ^146^.Water presaturation was conducted for 5 seconds during the relaxation period, followed by a spin-echo delay of 0.3 ms. Spectra were acquired at 298 K. The time-domain data underwent a weighted Fourier transform with 0.5 Hz line broadening. Manual adjustments for phase and baseline were made for profiling analysis. Resonance assignments and metabolites quantification was performed using Chenomx NMR Suite (version 8.6, Chenomx Inc., Edmonton, Alberta, Canada, https://www.chenomx.com) software, and data analysis was confirmed with Bayesil software ^144^.

### Statistical analysis of NMR data

The primary outcomes were serum metabolomic profiles and amino acid concentrations. The main exposures were PD subtype (iPD, gPD, rvPD) and sex. Covariates included age, disease duration, and L-DOPA equivalent daily dose (LEDD).

The matrices containing metabolite concentration data obtained from NMR peak quantifications were analysed using a univariate approach, combining a T-test with fold change analysis represented in a Robust volcano plot. A fold change threshold of 2 and a p-value threshold of less than 0.05 were set ^147^.

The matrices were normalised using sum and Pareto scaling before analysis. Partial least-squares discriminant analysis (PLS-DA) was performed on the normalised metabolomics data using MetaboAnalyst 6.0 (http://www.metaboanalyst.ca/) ^53^.

The PLS-DA model’s performance was assessed using the Q^2^ coefficient and accuracy, which employs a 10-fold internal cross-validation method ^148^. Both parameters are considered significant if they yield positive values.

Discriminatory metabolites were systematically organised and ranked according to their variable influence on projection (VIP) scores. The VIP scores, derived from the weighted sums of squares of the PLS-DA weights, indicate the importance of each variable and are considered statistically significant t when they exceed 1 ^149^.

We conducted pathway analysis with the Enrichment tool, referencing the Small Molecules Pathways Database (SMPDB) for Homo sapiens. Only pathways with a false discovery rate (FDR) and adjusted p-values below 0.05, along with more than one associated metabolite (hits), were included ^150^.

Analysis of biomarkers involved examining the univariate ROC curve to determine the AUC and its 95% confidence intervals (using 500 bootstrap cycles) ^53^. No missing data were present in the analysed variables.

### HPLC analysis of amino acids content

Serum samples (100 μl) were mixed in a 1:10 dilution with HPLC-grade methanol (900 μl) and centrifuged at 13,000 ×g for 10 min, as previously reported ^151^. Supernatants were dried and then suspended in 0.2 M trichloroacetic acid (TCA). TCA supernatants were then neutralized with 0.2 M NaOH and subjected to precolumn derivatization with o-phthaldialdehyde /N-acetyl-L-cysteine in 50% methanol. Amino acids derivatives were resolved on a UHPLC Nexera X3 system (Shimadzu) by using a Shimpack GIST C18 3-μm reversed-phase column (Shimadzu, 4.0 × 150 mm) under isocratic conditions (0.1 M sodium acetate buffer, pH 6.2, 1% tetrahydrofuran, and 1 mL/min flow rate). A washing step in 0.1 M sodium acetate buffer, 3% tetrahydrofuran, and 47% acetonitrile, was performed after every run. Identification and quantification of amino acids were based on retention times and peak areas and were compared with those of external standards. The detected amino acid concentration was expressed in μM, while the ratios were expressed as percentages.

### Statistical analysis of HPLC data

Normality distribution was assessed by q-q plot and Shapiro–Wilk test. Quantitative variables were expressed by the median and interquartile range (IQR), while qualitative variables were by absolute frequency. Differences between two independent groups were assessed using the non-parametric Mann–Whitney U test. Comparisons between more than two independent groups were performed using the non-parametric Kruskal–Wallis test, followed, when statistically significant, by Dunn’s post hoc test with Bonferroni correction for multiple comparisons. The potential confounding effects of age, LEDD and disease duration were assessed using analysis of covariance (ANCOVA). When variables exhibited non-normal distributions, natural logarithmic transformation was applied prior to analysis. The correlation of serum amino acid concentration with demographic and clinical parameters was evaluated with Spearman’s correlation test. Partial correlation analyses were adopted to test the effect of potential confounders. Significance was set at p < 0.05 for all analyses.

### Study cohorts adopted for genetic associations analysis

#### MNI-PD cohort

The Mediterranean Neurological Institute (MNI)-PD cohort includes 804 independent and unrelated PD patients (501 males; 300 familiar and 504 sporadic cases), for which Whole exome sequencing (WES) data are available. The MNI-PD cohort is part of the Parkinson’s disease Biobank of the IRCCS Neuromed and of the Institute of Genetics and Biophysics (IGB-CNR). The entire MNI cohort was used in case-control study to identify risk factors for PD. All the subjects were of European ancestry and were evaluated by qualified neurologists of the Parkinson Centre of the IRCCS INM Neuromed from June 2015 to December 2017, and from June 2021 to December 2023, with a thorough protocol comprising neurological examination and evaluation of non-motor domains. Information about family history, demographic characteristics, anamnesis, and pharmacological therapy was also collected (the treatment of the PD groups consisted for the most part of a combination of levodopa and dopamine agonists) ^15,16^.

#### MNI-rvPD cohort

This cohort was selected from the entire MNI-PD cohort of 804 PD patients with the same criteria adopted to select the PD cohort that we used in NMR/HPLC analyses. It includes 371 patients carrying at least one rare variant in PD genes (see the paragraph “Genetic stratification of the Study Cohort”). This cohort was used as validation cohort in genetic association analysis to confirm the association with PD in rvPD patients stratified by sex.

#### MNI-HC cohort

282 neurological healthy controls (HCs) were recruited by the same group of neurologists, among the patients’ wives/husbands, after having ascertained the lack of neurological pathologies and the absence of affected family members. WES data were available for the entire cohort. The MNI-HC cohort was used for association analysis.

#### Replication cohort

4,586 PD patients (from PDGC cohort) and 43,989 CNT (from UK biobank) whose data were downloaded from the PDGC Variant browser (https://pdgenetics.shinyapps.io/VariantBrowser/). For this dataset only aggregated genotypes were available regardless of sex and PD subtypes.

### Association analysis

Principal Component Analysis (PCA) was performed with Plink software and was used to characterize the genetic diversity of the study sample (PD_MNI, CNT_MNI) ^152^. The analysis was carried out by using common variants (Minor allele frequency MAF > 0.01), PC1 and PC2 were found to contribute to a variance of 25% among samples.

To identify the genetic contribution given by common variants (MAF > 0.01 referred to our internal database) we adopted a logistic regression model through plink2 software, by adjusting for age and the 10 principal components; we also adjusted for sex when we analyzed the entire cohort regardless of sex. p-values were adjusted for Bonferroni multiple testing correction.

### Expression studies in human brain regions

The Genotype-Tissue Expression (GTEx) portal (https://www.gtexportal.org/home/) was accessed to obtain gene expression data of the identified SNPs. The analysis was performed across all available adult human brain regions (amygdala, anterior cingulate cortex BA24, caudate nucleus, putamen, substantia nigra, cerebellar hemispheres, cerebellum, cerebral cortex, frontal cortex BA9, hippocampus, hypothalamus, nucleus accumbens).

## Declarations

## Data Availability

Metabolomics data have been deposited to the EMBL-EBI MetaboLights database (https://www.ebi.ac.uk/metabolights/) with the identifier **MTBLS12746**

## Code Availability

Not applicable

## Acknowledgments

The authors are grateful to all the patients, their caregivers, the Clinical Parkinson’s Disease Center of IRCCS Pozzilli and the PD biobank of the IRCCS Neuromed and of the IGB-CNR for the kind cooperation with this study.

This study was partially funded by Italian Ministry of University and Research (PRIN 2022 - COD. 2022XF7YYL to AU and PRIN 2022 – COD. 2022W3RKLJ to TE). The work of A.U., T.N. and T.E. was supported by NEXTGENERATIONEU (NGEU) and funded by the Ministry of University and Research (MUR), National Recovery and Resilience Plan (NRRP), project MNESYS (PE0000006) – A Multiscale integrated approach to the study of the nervous system in health and disease (DN. 1553 11.10.2022). The work of T.E. was supported by Next Generation EU - PNRR M6C2 Investimento 2.1 valorizzazione e potenziamento della ricerca biomedica del SSN grant n. PNRR-MAD-2022-12375960 and grant n. PNRR-MCNT2-2023-12377375. TE was also supported by Ministry of Health, Ricerca Corrente. The study of TE was partially funded by Ministry of Enterprises and Made in Italy (MIMIT) project Neurotechno n. F/180029/01/X43. The funders had no role in study design, data collection and analysis, decision to publish, or preparation of the manuscript.

## Author’s contributions

CM: Investigation, Data analysis, Writing –review & editing; FC: Data analysis, Writing –review & editing; TN: Data analysis, review & editing; MS: Writing –review & editing; IY: TN: Data analysis, review & editing; MG: Investigation, review & editing; SP: review & editing; NM: review & editing; FE: Writing – review & editing; AMDU: Supervision, review & editing; TE: Funding acquisition, Project administration, Supervision, Writing – review & editing; AU: Conceptualization, Funding acquisition, Project administration, Supervision, Writing – review & editing.

