## Supplementary data for "Discovery of Sexual Dimorphism in the Serum Metabolome of Parkinson’s Disease Patients Harboring Rare Genetic Variants with Uncertain Pathogenicity"

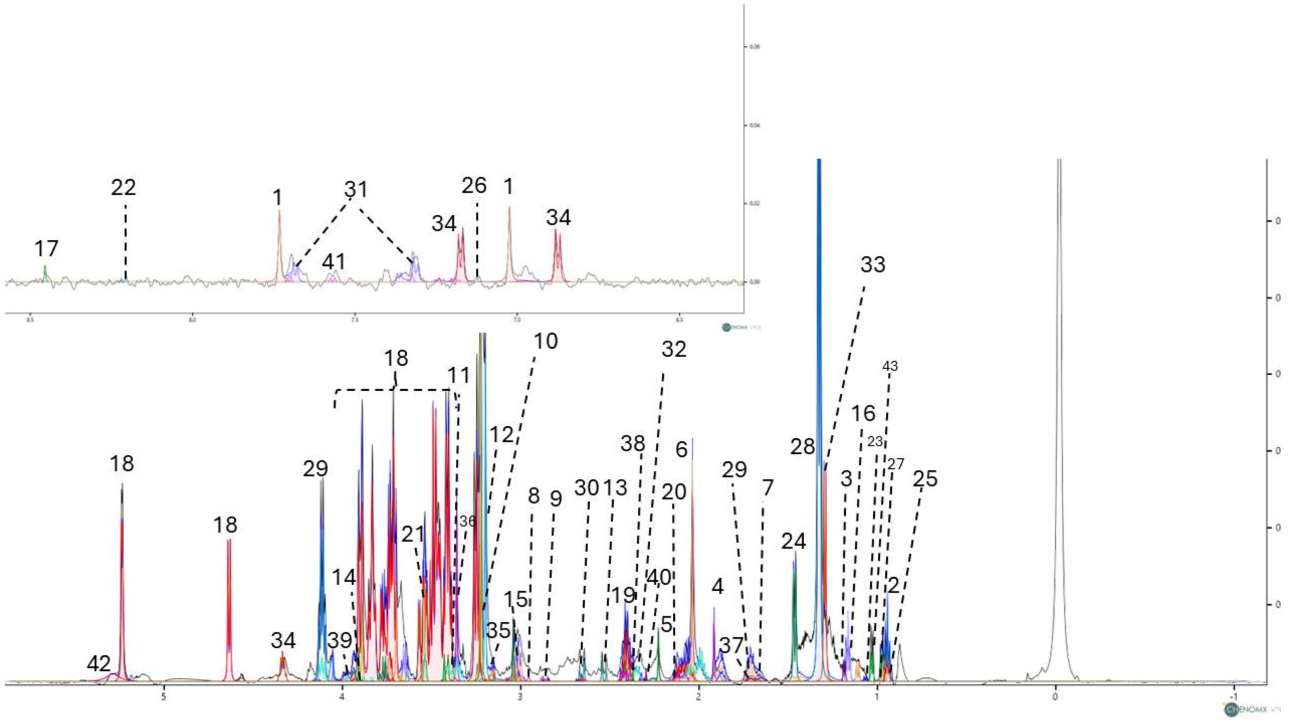


**Supplementary Figure 1. 1D-$^{1}$H-CPMG spectrum obtained from the serum of rvPD patients.** The spectrum is recorded at 600 MHz with a temperature of 298 K. A total of forty-three metabolites are identified and labelled as follows:1:1-Methylhistidine; 2:2-Hydroxybutyrate; 3:3-Hydroxybutyrate; 4:Acetic acid; 5:Acetoacetate; 6:Acetone; 7:L-Arginine; 8:L-Asparagine; 9:Aspartate; 10: Betaine; 11:Carnitine; 12:Choline; 13:Citric acid; 14:Creatine; 15:Creatinine; 16:Ethanol; 17:Formate; 18:D-Glucose; 19:L-Glutamic acid; 20:L-Glutamine; 21:Glycine; 22:Hypoxantine; 23:Isobutyrate; 24: L-Alanine; 25:L-Leucine; 26:L-Histidine; 27:Isoleucine; 28:L-Lactic acid; 29: L-Lysine; 30:Methionine; 31:L-Phenylalanine, 32:L-Proline; 33:L-Threonine; 34:Tyrosine; 35:Malonate; 36:Methanol; 37:L-Ornithine; 38:Pyruvic acid; 39: L-Serine, 40:Succinate; 41:Tryptophan; 42:Urea; 43:Valine.

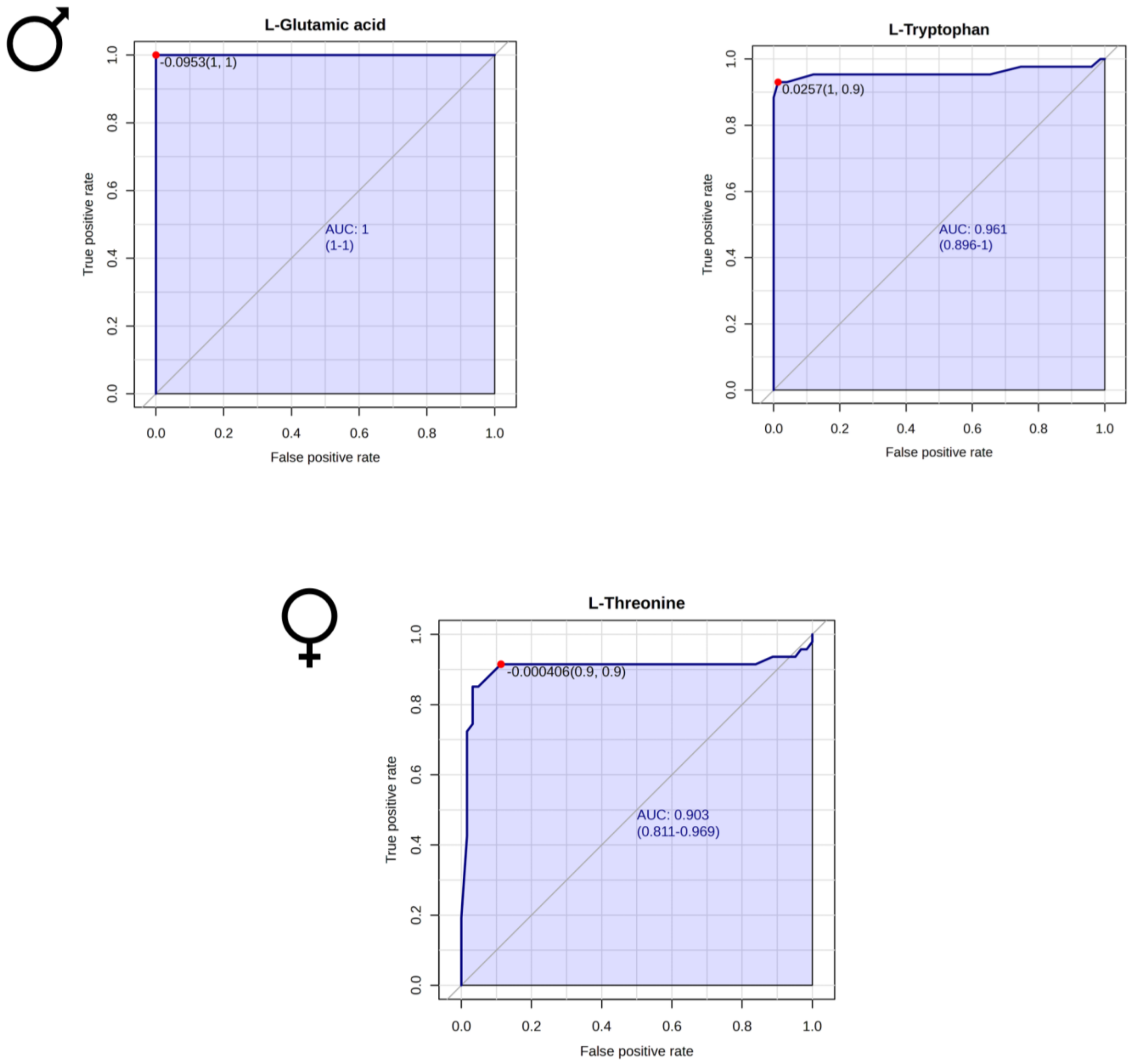


**Supplementary Figure 2. Receiver operating characteristic (ROC) curves for male and female rvPD patients compared to healthy controls (HC)**. The Cartesian space is described by the x-axis as the false positive rate and the y-axis as the true positive rate. The ROC curve comprises two components: the empirical ROC curve, obtained by connecting the points represented by sensitivity and specificity across various cut points, and the chance diagonal, represented by the 45-degree line drawn through the coordinates (0,0) and (1,1).

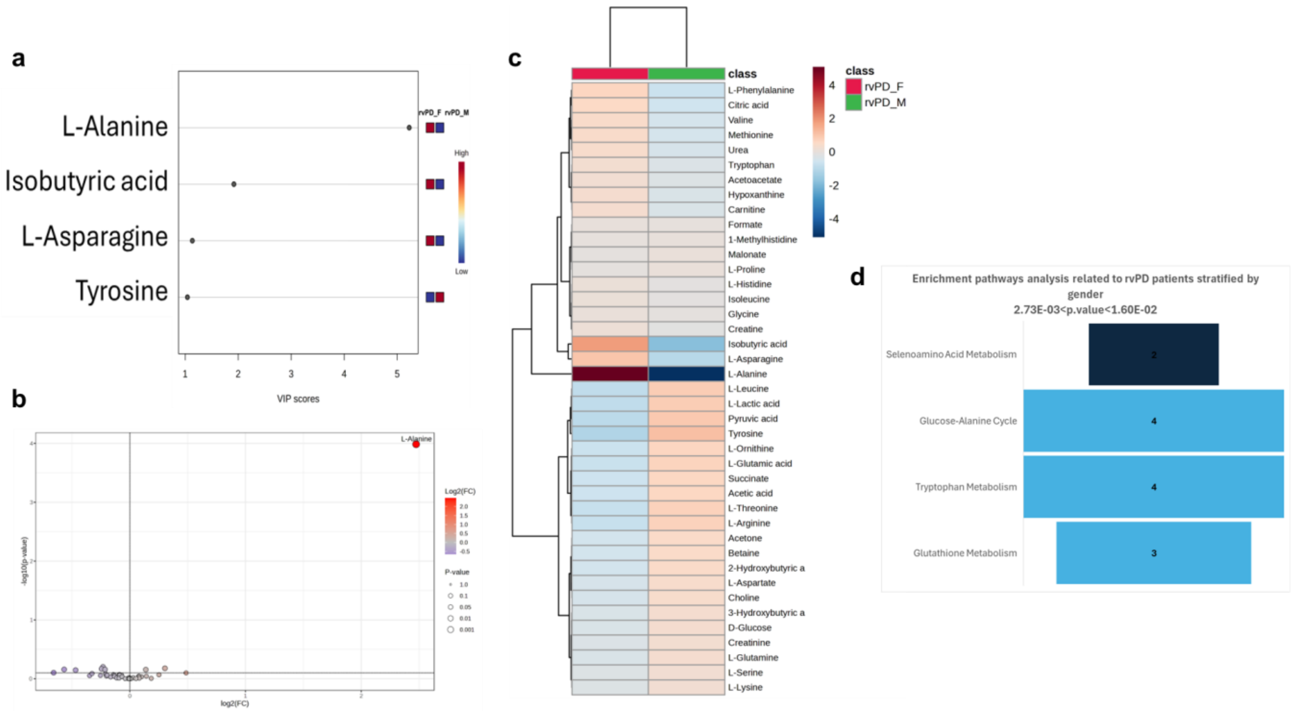


**Supplementary Figure 3. $^{1}$H-NMR-based metabolomic analysis reveals selective differences in serum profiles of rvPD stratified by sex a,** Variable Importance Projection plots reporting the metabolites responsible for rvPD male and female cluster separation. Only metabolites with VIP > 1 were considered significant. **b**, Robust volcano plots showing the upregulated and downregulated metabolites in the female rvPD serum, represented in red and blue, respectively. The fold-change value was set to 2, while the p-value threshold was set to <0.05. Metabolites significant for both fold-change and p-value from the t-test are labelled with an asterisk. **c,** Heatmaps of the modified metabolites relative to rvPD patients. The colour of each section corresponds to the concentration value of each metabolite, calculated from a normalised concentration matrix (red, upregulated; blue, downregulated). **d**, Pathway enrichment analysis was performed on $^{1}$H-NMR data related to serum metabolomics results of rvPD patients stratified for sex. The bars represent the hits, i.e., the number of metabolites detected in the spectra and involved in the pathways. Pathways were considered statistically significant with hits > 1, p-value < 0.05, and p-values adjusted using the Holm–Bonferroni test (Holm p) and the False Discovery Rate (FDR) < 1. Darker colours represent lower and, therefore, more significant p-values. The range of significance is shown at the top. The exploration of pathways was carried out using the Small Molecule Pathway Database (SMPDB), selecting *Homo sapiens* as the organism.

**Supplementary Table 1.** Rare (MAF≤0.01) and pathogenic variants identified in the rvPD and gPD cohort**.**

| CHR | Genomic position (hg38) | dbSNP | Gene | RefSEq | Nucl Change | AA Change | EF | CADD phred | MAF* | N. PD patients |
|---|---|---|---|---|---|---|---|---|---|---|
| 7 | 6009379 | rs139842556 | AIMP2 | NM_006303 | c.G16C | p.V6L | NSV | 25.8 | NA | 1 |
| 7 | 6017994 | rs542587504 | AIMP2 | NM_006303 | c.A523G | p.K175E | NSV | 13.30 | NA | 1 |
| 7 | 6023405 | rs139398981 | AIMP2 | NM_006303 | c.A677G | p.N226S | NSV | 16.35 | 0.00039 | 1 |
| 11 | 113388065 | rs181883517 | ANKK1 | NM_178510 | c.G181A | p.A61T | NSV | 0.082 | 0.00019 | 2 |
| 11 | 113393582 | NA | ANKK1 | NM_178510 | c.T287G | p.I96S | NSV | 24.9 | NA | 1 |
| 11 | 113393583 | NA | ANKK1 | NM_178510 | c.T288G | p.I96M | NSV | 10.50 | NA | 1 |
| 11 | 113395070 | rs770164219 | ANKK1 | NM_178510 | c.G622T | p.D208Y | NSV | 23.3 | NA | 1 |
| 11 | 113395381 | NA | ANKK1 | NM_178510 | c.C655A | p.L219I | NSV | 1.136 | NA | 1 |
| 11 | 113397225 | rs138608171 | ANKK1 | NM_178510 | c.C840A | p.D280E | NSV | 10.88 | 0.00099 | 1 |
| 11 | 113399756 | rs7104979 | ANKK1 | NM_178510 | c.C1787T | p.P596L | NSV | 23.2 | 0.00499 | 2 |
| 4 | 124669880 | NA | ANKRD50 | NM_020337 | c.A3397G | p.I1133V | NSV | 17.99 | NA | 1 |
| 4 | 124671095 | rs377077898 | ANKRD50 | NM_020337 | c.G2182A | p.A728T | NSV | 20.5 | 7.5e-05 | 1 |
| 4 | 124672177 | rs140232140 | ANKRD50 | NM_020337 | c.C1100T | p.T367M | NSV | 17.50 | 0.0059 | 2 |
| 3 | 132463816 | NA | DNAJC13 | NM_020337 | c.A1891G | p.R631G | NSV | 29.4 | NA | 1 |
| 3 | 132467276 | NA | DNAJC13 | NM_020337 | c.T2171C | p.M724T | NSV | 20.3 | NA | 1 |
| 3 | 132475009 | rs149121829 | DNAJC13 | NM_020337 | c.C2369A | p.S790Y | NSV | 23.3 | 0.00439 | 1 |
| 3 | 132477831 | rs752266405 | DNAJC13 | NM_020337 | c.T2488C | p.Y830H | NSV | 25.2 | NA | 1 |
| 3 | 132478139 | rs141952333 | DNAJC13 | NM_020337 | c.G2708A | p.R903K | NSV | 26.3 | 0.00139 | 1 |
| 3 | 132480382 | NA | DNAJC13 | NM_020337 | c.A2786G | p.D929G | NSV | 24.5 | NA | 1 |
| 3 | 132499231 | rs61748102 | DNAJC13 | NM_020337 | c.C4262T | p.A1421V | NSV | 23.9 | 0.01277 | 1 |

| | | | | | | | | | | |
|---|---|---|---|---|---|---|---|---|---|---|
| 3 | 132499777 | rs61748103 | DNAJC13 | NM_015575 | c.G4385A | p.R1462H | NSV | 22.6 | 0.00559 | 1 |
| 3 | 132502349 | rs759696878 | DNAJC13 | NM_015575 | c.T4597C | p.W1533R | NSV | 23.2 | NA | 1 |
| 3 | 132522838 | rs145242123 | DNAJC13 | NM_015575 | c.C5684T | p.T1895M | NSV | 16.68 | 0.00059 | 2 |
| 3 | 132523547 | rs569025407 | DNAJC13 | NM_015575 | c.A5894G | p.E1965G | NSV | 23.6 | 0.00019 | 1 |
| 3 | 132525682 | rs145794875 | DNAJC13 | NM_015575 | c.G6133A | p.V2045I | NSV | 20.4 | NA | 2 |
| 3 | 132528264 | rs748912838 | DNAJC13 | NM_015575 | c.G6457A | p.A2153T | NSV | 22.6 | NA | 1 |
| 1 | 65392454 | rs145329294 | DNAJC6 | NM_001256864 | c.T1492A | p.C498S | NSV | 9.480 | 0.00279 | 2 |
| 1 | 65392476 | NA | DNAJC6 | NM_001256864 | c.T1514C | p.L505P | NSV | 23.7 | NA | 1 |
| 3 | 184321366 | rs774842696 | EIF4G1 | NM_182917 | c.C782T | p.S261L | NSV | 21.1 | NA | 1 |
| 3 | 184321452 | rs974387728 | EIF4G1 | NM_182917 | c.A868G | p.M290V | NSV | 0.067 | NA | 1 |
| 3 | 184322040 | rs112545306 | EIF4G1 | NM_182917 | c.C1456T | p.P486S | NSV | 0.111 | 0.00019 | 1 |
| 3 | 184322583 | rs111924994 | EIF4G1 | NM_182917 | c.G1648C | p.A550P | NSV | 13.88 | 0.00119 | 1 |
| 3 | 184322602 | rs772950492 | EIF4G1 | NM_182917 | c.C1667T | p.P556L | NSV | 19.78 | NA | 1 |
| 3 | 184327401 | rs112176450 | EIF4G1 | NM_182917 | c.G3617A | p.R1206H | NSV | 24.9 | NA | 1 |
| 3 | 184327433 | rs201711322 | EIF4G1 | NM_182917 | c.C3649T | p.R1217C | NSV | 30 | NA | 1 |
| 3 | 184327609 | rs35629949 | EIF4G1 | NM_182917 | c.C3688G | p.P1230A | NSV | 0.346 | 0.00139 | 1 |
| 3 | 184328723 | rs765574482 | EIF4G1 | NM_182917 | c.A4049G | p.Q1350R | NSV | 22.3 | NA | 2 |
| 22 | 32479013 | rs550610502 | FBXO7 | NM_012179 | c.A155G | p.Y52C | NSV | 24.0 | 0.00119 | 1 |
| 22 | 32479102 | rs757307611 | FBXO7 | NM_012179 | c.A244G | p.I82V | NSV | 17.89 | NA | 1 |
| 22 | 32484000 | rs376455464 | FBXO7 | NM_012179 | c.C521T | p.S174L | NSV | 22.1 | NA | 1 |
| 22 | 32498507 | rs34316445 | FBXO7 | NM_012179 | c.G1546T | p.D516Y | NSV | 25.0 | NA | 1 |
| **1** | **155235843** | **rs76763715** | **GBA1** | **NM_000157** | **c.A1226G** | **p.N409S** | **NSV** | **24.1** | **0.001728** | **9** |
| **1** | **155236376** | **rs2230288** | **GBA1** | **NM_000157** | **c.G1093A** | **p.E365K** | **NSV** | **16.1** | **0.01338** | **7** |

| | | | | | | | | | | |
|---|---|---|---|---|---|---|---|---|---|---|
| **1** | **155236246** | **rs75548401** | **GBA1** | **NM_000157** | **c.C1223T** | **p.T408M** | **NSV** | **21.3** | **0.008175** | **3** |
| **1** | **155235252** | **rs421016** | **GBA1** | **NM_000157** | **c.T1448C** | **p.L483P** | **NSV** | **24.7** | **0.00007547** | **2** |
| **1** | **155237458** | **rs367968666** | **GBA1** | **NM_000157** | **c.T882G** | **p.H294Q** | **NSV** | **12.25** | **0.0001678** | **2** |
| **1** | **155240696** | **rs139626710** | **GBA1** | **NM_000157** | **c.A49G** | **p.R17G** | **NSV** | **8.03** | **0.00002119** | **1** |
| **1** | **155239995** | **NA** | **GBA1** | **NM_000157** | **c.C198A** | **p.D66E** | **NSV** | **6.32** | **NA** | **1** |
| **1** | **155238206** | **rs381427** | **GBA1** | **NM_000157** | **c.T689A** | **p.V230E** | **NSV** | **22.3** | **0.00000339** | **1** |
| **1** | **155238177** | **NA** | **GBA1** | **NM_000157** | **c.C718T** | **p.P240S** | **NSV** | **22.4** | **NA** | **1** |
| **1** | **155236370** | **NA** | **GBA1** | **NM_000157** | **c.C1099T** | **p.H367Y** | **NSV** | **25.7** | **NA** | **1** |
| **1** | **155236261** | **rs121908307** | **GBA1** | **NM_000157** | **c.G1208C** | **p.S403T** | **NSV** | **22.8** | **8.476e-7** | **1** |
| **1** | **155235790** | **rs149171124** | **GBA1** | **NM_000157** | **c.G1279A** | **p.E427K** | **NSV** | **19.29** | **0.0002229** | **1** |
| **1** | **155235727** | **rs1064651** | **GBA1** | **NM_000157** | **c.G1342C** | **p.D448H** | **NSV** | **23.3** | **0.0001043** | **1** |
| **1** | **155235205** | **rs369068553** | **GBA1** | **NM_000157** | **c.G1495C** | **p.V499L** | **NSV** | **17.25** | **0.00004747** | **1** |
| **1** | **155235091** | **NA** | **GBA1** | **NM_000157** | **c.G1515T** | **p.K505N** | **NSV** | **19.17** | **NA** | **1** |
| 2 | 232760519 | NA | GIGYF2 | NM_015575 | c.T419A | p.F140Y | NSV | 27.4 | NA | 1 |
| 2 | 232832931 | rs369380236 | GIGYF2 | NM_015575 | c.G2604A | p.M868I | NSV | 22.8 | NA | 1 |
| 2 | 232839964 | rs146430802 | GIGYF2 | NM_015575 | c.G2882A | p.R961Q | NSV | 25.3 | 0.00059 | 1 |
| 2 | 232847553 | rs763368000 | GIGYF2 | NM_015575 | c.3666_3677del | p.P1225_Q1228del | NFDel | NA | NA | 3 |
| 19 | 14479503 | rs147583484 | GIPC1 | NM_005716 | c.C677T | p.A226V | NSV | 2.377 | 0.00199 | 1 |
| 16 | 4507845 | rs150288371 | HMOX2 | NM_001286271 | c.A250T | p.M84L | NSV | 14.00 | 0.00019 | 3 |
| 16 | 4509415 | rs145119833 | HMOX2 | NM_001286271 | c.T613C | p.F205L | NSV | 24.7 | NA | 1 |
| 16 | 4509433 | rs149714752 | HMOX2 | NM_001286271 | c.G631C | p.A211P | NSV | 23.9 | 0.00099 | 1 |
| 16 | 4509709 | rs375786109 | HMOX2 | NM_001286271 | c.G817A | p.A273T | NSV | 7.929 | NA | 1 |

| | | | | | | | | | | |
|---|---|---|---|---|---|---|---|---|---|---|
| 11 | 123059263 | NA | HSPA8 | NM_006597 | c.1121-2->T | NA | splicing | NA | 8,00E-05 | 2 |
| 2 | 74531576 | NA | HTRA2 | NM_001321727 | c.C949A | p.L317M | NSV | 13.03 | NA | 1 |
| 2 | 74532698 | rs72470545 | HTRA2 | NM_013247 | c.G1195A | p.G399S | NSV | 26.9 | 0.00359 | 7 |
| 2 | 86144339 | NA | IMMT | NM_001100169 | c.C2203G | p.Q735E | NSV | 26.5 | NA | 1 |
| 2 | 86144516 | NA | IMMT | NM_001100169 | c.C2026T | p.P676S | NSV | 24.3 | NA | 1 |
| 2 | 86151395 | rs61731709 | IMMT | NM_001100169 | c.G1300A | p.E434K | NSV | 26.6 | 0.00119 | 1 |
| 2 | 86171288 | rs368629905 | IMMT | NM_001100169 | c.456_479del | p.G153_A160del | NFDel | NA | NA | 1 |
| 1 | 200988535 | NA | KIF21B | NM_001252100 | c.A3308G | p.Y1103C | NSV | 22.4 | NA | 1 |
| 1 | 200990930 | rs760098433 | KIF21B | NM_001252100 | c.A2674T | p.T892S | NSV | 13.36 | NA | 1 |
| 1 | 201000528 | NA | KIF21B | NM_001252100 | c.C1547G | p.S516C | NSV | 22.2 | NA | 1 |
| 1 | 201008899 | rs140589352 | KIF21B | NM_001252100 | c.C317T | p.S106L | NSV | 18.73 | 0.00838 | 5 |
| 9 | 34256237 | rs145898456 | KIF24 | NM_194313 | c.G3370A | p.G1124S | NSV | 9.330 | 0.000199 | 2 |
| 9 | 34256377 | rs34101674 | KIF24 | NM_194313 | c.C3230A | p.T1077K | NSV | 0.080 | 0.00559 | 5 |
| 9 | 34256581 | rs117856906 | KIF24 | NM_194313 | c.G3026A | p.R1009K | NSV | 0.564 | 0.00059 | 1 |
| 9 | 34256936 | rs148278926 | KIF24 | NM_194313 | c.A2671T | p.S891C | NSV | 22.7 | 0.00019 | 1 |
| 9 | 34269346 | rs752724092 | KIF24 | NM_194313 | c.T1354G | p.L452V | NSV | 23.7 | NA | 1 |
| 12 | 40235634 | rs33995463 | LRRK2 | NM_198578 | c.T356C | p.L119P | NSV | 27.4 | 0.00079 | 2 |
| 12 | 40249924 | NA | LRRK2 | NM_198578 | c.C937T | p.L313F | NSV | 24.2 | NA | 1 |
| 12 | 40252960 | NA | LRRK2 | NM_198578 | c.C1232G | p.S411X | stopgain | 35 | NA | 1 |
| 12 | 40263797 | rs770721531 | LRRK2 | NM_198578 | c.G1552A | p.E518K | NSV | 22.1 | NA | 1 |
| 12 | 40293613 | rs201184634 | LRRK2 | NM_198578 | c.G2758A | p.A920T | NSV | 2.812 | NA | 1 |
| 12 | 40304044 | NA | LRRK2 | NM_198578 | c.T3687A | p.H1229Q | NSV | 8.023 | NA | 1 |
| 12 | 40305791 | rs4640000 | LRRK2 | NM_198578 | c.C3784G | p.P1262A | NSV | 23.8 | 0.00878 | 1 |

| | | | | | | | | | | |
|---|---|---|---|---|---|---|---|---|---|---|
| 12 | 40308618 | rs17466213 | LRRK2 | NM_198578 | c.A4111G | p.I1371V | NSV | 20.2 | 0.00119 | 1 |
| 12 | 40320094 | rs759886320 | LRRK2 | NM_198578 | c.A4934G | p.Y1645C | NSV | 28.4 | NA | 1 |
| 12 | 40320097 | rs35303786 | LRRK2 | NM_198578 | c.T4937C | p.M1646T | NSV | 21.8 | 0.00479 | 5 |
| 12 | 40323256 | rs35602796 | LRRK2 | NM_198578 | c.T5606C | p.M1869T | NSV | 22.2 | 0.00019 | 1 |
| 12 | 40359345 | rs200002022 | LRRK2 | NM_198578 | c.C6929T | p.T2310M | NSV | 6.324 | NA | 2 |
| 12 | 40363401 | NA | LRRK2 | NM_198578 | c.7029-1G>T | NA | splicing | 34 | NA | 1 |
| 12 | 40367045 | rs146428335 | LRRK2 | NM_198578 | c.G7430A | p.R2477Q | NSV | 15.40 | 0.00019 | 1 |
| **12** | **40734202** | **rs34637584** | **LRRK2** | **NM_198578** | **c.G6055A** | **p.G2019S** | **NSV** | **35** | **0.0002721** | **14** |
| **12** | **40704236** | **rs33939927** | **LRRK2** | **NM_198578** | **c.C4321T** | **p.R1441C** | **NSV** | **26.7** | **0.0000195** | **2** |
| **12** | **40252984** | **rs34594498** | **LRRK2** | **NM_198578** | **c.C1256T** | **p.A419V** | **NSV** | **24.9** | **0.0001493** | **1** |
| 15 | 75356218 | rs754290709 | MAN2C1 | NM_001256494 | c.C2939T | p.A980V | NSV | 31 | NA | 1 |
| 15 | 75356347 | rs775890577 | MAN2C1 | NM_001256494 | c.C2891T | p.A964V | NSV | 23.2 | NA | 1 |
| 15 | 75356614 | rs769808322 | MAN2C1 | NM_001256494 | c.C2780T | p.P927L | NSV | 25.7 | NA | 1 |
| 15 | 75358481 | rs143005170 | MAN2C1 | NM_001256494 | c.A2435G | p.Y812C | NSV | 24.6 | 0.00059 | 1 |
| 15 | 75358562 | rs62029711 | MAN2C1 | NM_001256494 | c.G2354A | p.R785Q | NSV | 23.7 | 0.00219 | 4 |
| 15 | 75358753 | rs146675988 | MAN2C1 | NM_001256494 | c.C2248T | p.P750S | NSV | 26.6 | NA | 3 |
| 15 | 75361381 | NA | MAN2C1 | NM_001256494 | c.C1219G | p.H407D | NSV | 31 | NA | 1 |
| 15 | 75362344 | rs79908208 | MAN2C1 | NM_001256494 | c.T1007C | p.M336T | NSV | 24.7 | 0.0061 | 1 |
| 15 | 75364182 | rs190692217 | MAN2C1 | NM_001256494 | c.G607A | p.G203R | NSV | 18.00 | 0.00019 | 2 |
| 15 | 75368504 | rs760253802 | MAN2C1 | NM_001256494 | c.76_80del | p.F26Rfs*3 | FDel | NA | NA | 2 |
| 6 | 34528879 | NA | PACSIN1 | NM_020804 | c.456+2T>G | NA | splicing | 34 | NA | 2 |
| 6 | 162201207 | rs55654276 | PRKN | NM_004562 | c.C458G | p.P153R | NSV | 21.9 | 0.00638 | 1 |
| **6** | **161350139** | **rs775091228** | **PRKN** | **NM_004562** | **c.G1358A** | **p.W453X** | **Stop_gain** | **44** | **0.00001186** | 2 |

| | | | | | | | | | | |
|---|---|---|---|---|---|---|---|---|---|---|
| 6 | 161785820 | rs34424986 | PRKN | NM_004562 | c.C823T | p.R275W | NSV | 26.1 | 0.003732 | 2 |
| 6 | 161360169 | rs55830907 | PRKN | NM_004562 | c.C1204T | p.R402C | NSV | 25.1 | 0.001937 | 4 |
| 6 | 162262692 | rs55774500 | PRKN | NM_004562 | c.C245A | p.A82E | NSV | 3.14 | 0.002815 | 5 |
| 6 | 162054135 | rs9456735 | PRKN | NM_004562 | c.A574C | p.M192L | NSV | 20.7 | 0.0001356 | 1 |
| 6 | 161973335 | rs144032774 | PRKN | NM_004562 | c.G701A | p.R234Q | NSV | 20.9 | 0.0001280 | 1 |
| 6 | 161973317 | rs137853054 | PRKN | NM_004562 | c.C719T | p.T240M | NSV | 23.4 | 0.0001802 | 1 |
| 6 | 161973306 | NA | PRKN | NM_004562 | c.G730C | p.V244L | NSV | 6.93 | NA | 1 |
| 6 | 161360148 | NA | PRKN | NM_004562 | c.G1225T | p.E409X | Stop_gain | 40 | NA | 1 |
| 6 | 161360129 | rs778125254 | PRKN | NM_004562 | c.C1244A | p.T415N | NSV | 25.2 | 0.00000339 | 1 |
| 1 | 20638041 | rs138302371 | PINK1 | NM_032409 | c.C587T | p.P196L | NSV | 17.46 | 0.0003610 | 2 |
| 1 | 20649054 | rs74315356 | PINK1 | NM_032409 | c.G1311A | p.W437X | Stop_gain | 47 | 0.00000677 | 2 |
| 1 | 20637956 | rs768091663 | PINK1 | NM_032409 | c.G502C | p.A168P | NSV | 22.2 | 0.00000847 | 1 |
| 1 | 20638012 | rs143204084 | PINK1 | NM_032409 | c.G558C | p.K186N | NSV | 14.84 | 0.0002568 | 1 |
| 1 | 20644585 | NA | PINK1 | NM_032409 | c.C872A | p.A291D | NSV | 26.3 | NA | 1 |
| 1 | 20645576 | rs376323248 | PINK1 | NM_032409 | c.C976T | p.R326C | NSV | 26.9 | 0.00000508 | 1 |
| 1 | 20650518 | rs531477772 | PINK1 | NM_032409 | c.G1573A | p.D525N | NSV | 23.3 | 0.00009152 | 1 |
| 1 | 7970934 | rs71653619 | PARK7 | NM_007262 | c.G293A | p.R98Q | NSV | 20.9 | 0.01043 | 14 |
| 1 | 7969406 | rs781094807 | PARK7 | NM_007262 | c.252+2->A | | splicing | NA | 0.0002733 | 2 |
| 16 | 670326 | NA | RHOT2 | NM_138769 | c.G407A | p.S136N | NSV | 10.42 | NA | 1 |
| 16 | 670532 | rs768236479 | RHOT2 | NM_138769 | c.C515G | p.P172R | NSV | 24.1 | NA | 1 |
| 16 | 670965 | rs1045709 | RHOT2 | NM_138769 | c.C713T | p.A238V | NSV | 8.32 | 0.00139 | 1 |
| 16 | 671067 | rs200255907 | RHOT2 | NM_001352287 | c.C70T | p.L24F | NSV | NA | 0.00019 | 1 |

| | | | | | | | | | | |
|---|---|---|---|---|---|---|---|---|---|---|
| 16 | 672522 | rs142157910 | RHOT2 | NM_138769 | c.G1360A | p.A454T | NSV | 19.28 | 0.00039 | 1 |
| 16 | 673506 | rs147973504 | RHOT2 | NM_138769 | c.C1757T | p.P586L | NSV | 11.86 | 0.00279 | 5 |
| 16 | 673545 | NA | RHOT2 | NM_138769 | c.1796_1797insGGCCGCCGT | p.V605_L606insAAV | NFIns | NA | NA | 1 |
| 17 | 44321168 | rs11545312 | SLC25A39 | NM_001143780 | c.T581G | p.V194G | NSV | 23.6 | NA | 1 |
| 17 | 44321169 | rs775602410 | SLC25A39 | NM_001143780 | c.G580A | p.V194M | NSV | 18.44 | NA | 1 |
| 17 | 44321703 | rs146901450 | SLC25A39 | NM_001143780 | c.C389T | p.T130I | NSV | 25.6 | 0.00039 | 1 |
| 5 | 1394741 | rs200712598 | SLC6A3 | NM_001044 | c.G1857C | p.K619N | NSV | 16.13 | 0.00039 | 1 |
| 5 | 1400949 | rs147837176 | SLC6A3 | NM_001044 | c.A1805G | p.E602G | NSV | 14.08 | 0.000399 | 1 |
| 5 | 1441392 | rs780852886 | SLC6A3 | NM_001044 | c.G385A | pA129T | NSV | 23.8 | NA | 1 |
| 5 | 1443117 | rs915327713 | SLC6A3 | NM_001044 | c.G81C | p.K27N | NSV | 23.1 | NA | 1 |
| 5 | 122401050 | NA | SNCAIP | NM_001308100 | c.C122T | p.A41V | NSV | 0.028 | NA | 1 |
| 5 | 122403816 | rs149915358 | SNCAIP | NM_001308106 | c.G29A | p.R10Q | NSV | 10.69 | 0.00439 | 4 |
| 5 | 122403846 | rs10070158 | SNCAIP | NM_005460 | c.G59A | p.R20H | NSV | 0.351 | 0.000798 | 2 |
| 5 | 122423128 | NA | SNCAIP | NM_005460 | c.G391A | p.G131S | NSV | 7.271 | NA | 1 |
| 5 | 122425422 | rs371777182 | SNCAIP | NM_005460 | c.C1073T | p.A358V | NSV | 24.3 | 0.000199 | 1 |
| 5 | 122425526 | rs778867427 | SNCAIP | NM_005460 | c.A1177G | p.I393V | NSV | 16.87 | NA | 1 |
| 5 | 122450664 | rs144492699 | SNCAIP | NM_005460 | c.G1817A | p.R606Q | NSV | 24.0 | NA | 1 |
| 5 | 122450972 | rs55712196 | SNCAIP | NM_005460 | c.G2125C | p.E709Q | NSV | 24.6 | 0.00519 | 4 |
| 5 | 122451264 | rs140850272 | SNCAIP | NM_005460 | c.G2417A | p.R806H | NSV | 28.5 | 0.00638 | 2 |
| 5 | 122451380 | rs758647740 | SNCAIP | NM_005460 | c.2533_2535del | p.Q845del | NSV | NA | NA | 1 |
| 5 | 122451491 | NA | SNCAIP | NM_005460 | c.C2644T | p.Q882X | stopgain | 37 | NA | 1 |

| | | | | | | | | | | |
|---|---|---|---|---|---|---|---|---|---|---|
| 5 | 122451557 | rs751412863 | SNCAIP | NM_005460 | c.A2710G | p.K904E | NSV | 25.5 | NA | 1 |
| 5 | 122463520 | NA | SNCAIP | NM_001242935 | c.A1757T | p.K586I | NSV | 26.2 | NA | 1 |
| 5 | 122463542 | rs187197845 | SNCAIP | NM_001242935 | c.T1779G | p.I593M | NSV | 15.27 | 0.00119 | 4 |
| 2 | 54618118 | rs760099132 | SPTBN1 | NM_003128 | c.G688A | p.A230T | NSV | 27.4 | NA | 1 |
| 2 | 54626143 | rs201568567 | SPTBN1 | NM_003128 | c.G1553A | p.R518K | NSV | 14.54 | 0.000399 | 1 |
| **4** | **952418** | **rs752406714** | **TMEM175** | **NM_032326** | **c.430_431del** | **p.V147Dfs*104** | **Fs_del** | **NA** | **0.00000256** | **4** |
| **4** | **958262** | **rs565504915** | **TMEM175** | **NM_032326** | **c.1281_1282del** | **p.A429Qfs*120** | **Fs_del** | **NA** | **0.0003095** | **3** |
| **4** | **952434** | **NA** | **TMEM175** | **NM_032326** | **c.446_462del** | **p.A149Gfs*97** | **Fs_del** | **NA** | **NA** | **3** |
| **4** | **947842** | **rs542936413** | **TMEM175** | **NM_032326** | **c.C103T** | **p.R35C** | **NSV** | **28.7** | **0.00002542** | **3** |
| **4** | **958194** | **rs75307864** | **TMEM175** | **NM_032326** | **c.C1213G** | **p.L405V** | **NSV** | **23.3** | **0.003832** | **4** |
| **4** | **958221** | **rs140597786** | **TMEM175** | **NM_032326** | **c.C1240T** | **p.R414W** | **NSV** | **23.2** | **0.004833** | **3** |
| **4** | **957985** | **rs147762522** | **TMEM175** | **NM_032326** | **c.G1004A** | **p.R335H** | **NSV** | **24.6** | **0.001076** | **3** |
| **4** | **950461** | **rs142778595** | **TMEM175** | **NM_032326** | **c.T233C** | **p.I78T** | **NSV** | **23.4** | **0.00005424** | **2** |
| **4** | **951229** | **rs200834686** | **TMEM175** | **NM_032326** | **c.A313G** | **p.T105A** | **NSV** | **22.21** | **0.0001136** | **2** |
| **4** | **955826** | **rs746980739** | **TMEM175** | **NM_032326** | **c.C778T** | **p.R260C** | **NSV** | **25.1** | **0.00004068** | **1** |
| **4** | **955856** | **rs750645874** | **TMEM175** | **NM_032326** | **c.G808A** | **p.A270T** | **NSV** | **25.0** | **0.00000762** | **1** |
| **4** | **955888** | **NA** | **TMEM175** | **NM_032326** | **c.C840G** | **p.I280M** | **NSV** | **24.7** | **NA** | **1** |
| **4** | **957838** | **rs778399444** | **TMEM175** | **NM_032326** | **c.C857T** | **p.P286L** | **NSV** | **23.9** | **0.00000254** | **1** |
| **4** | **957958** | **rs148627215** | **TMEM175** | **NM_032326** | **c.C977T** | **p.A326V** | **NSV** | **7.41** | **0.00000169** | **1** |
| **4** | **958024** | **rs147975675** | **TMEM175** | **NM_032326** | **c.C1043T** | **p.S348L** | **NSV** | **24.5** | **0.00005171** | **1** |
| **4** | **958252** | **rs142744759** | **TMEM175** | **NM_032326** | **c.C1271T** | **p.A424V** | **NSV** | **22.6** | **0.00001188** | **1** |
| **4** | **958422** | **rs201314478** | **TMEM175** | **NM_032326** | **c.C1441T** | **p.R481W** | **NSV** | **10.6** | **0.002678** | **1** |

| | | | | | | | | | | |
|---|---|---|---|---|---|---|---|---|---|---|
| 16 | 10770298 | rs371043030 | TVP23A | NM_001079512 | c.G616A | p.E206K | NSV | 8.519 | 0.000199 | 3 |
| 4 | 41261901 | NA | UCHL1 | NM_004181 | c.T437G | p.V146G | NSV | 32 | NA | 1 |
| 16 | 46672341 | rs139200616 | VPS35 | NM_018206 | c.A1292T | p.E431V | NSV | 23.3 | NA | 1 |
| 16 | 46672456 | rs779224141 | VPS35 | NM_018206 | c.1176_1177insAGT | p.S392_A393insS | NFIns | NA | NA | 1 |
| 3 | 184824692 | rs192763506 | VPS8 | NM_015303 | c.A60T | p.E20D | NSV | 8.300 | 0.00139 | 1 |
| 3 | 184855772 | NA | VPS8 | NM_015303 | c.A1091T | p.N364I | NSV | 25.9 | NA | 1 |
| 3 | 184886111 | rs746576786 | VPS8 | NM_015303 | c.A1730G | p.D577G | NSV | 27.5 | NA | 1 |
| 3 | 184886122 | rs61742617 | VPS8 | NM_015303 | c.G1741A | p.V581I | NSV | 17.35 | 0.000998 | 3 |
| 3 | 184894756 | rs201387135 | VPS8 | NM_015303 | c.A1829G | p.K610R | NSV | 22.0 | NA | 1 |
| 3 | 184966686 | rs527847853 | VPS8 | NM_015303 | c.C3283T | p.L1095F | NSV | 22.9 | 0.000199 | 1 |
| 3 | 185048537 | rs16859527 | VPS8 | NM_015303 | c.G4109A | p.R1370H | NSV | 16.97 | 0.0115 | 1 |
| 7 | 100057239 | rs147509446 | ZSCAN21 | NM_145914 | c.T233G | p.I78S | NSV | 24.2 | 0.00079 | 1 |
| 7 | 100057743 | rs778353607 | ZSCAN21 | NM_145914 | c.T445C | p.S149P | NSV | 9.891 | NA | 1 |
| 7 | 100063991 | rs146827623 | ZSCAN21 | NM_145914 | c.G796A | p.V266I | NSV | 2.470 | 0.000199 | 1 |
| 7 | 100064601 | rs776436945 | ZSCAN21 | NM_145914 | c.A1406G | p.E469G | NSV | 26.9 | NA | 1 |

CHR, Chromosome; hg38, human genome build to which these variants are annotated; dbSNP, reference number in SNP database; NA, Not Annotated; ref seq, reference number of the gene transcript; Nucl Change, nucleotide change; AA Change, amino acid change; EF, Exonic Function; FDel, Frame shift deletion; NFIns, Non-Frame shift insertion; NFDel, Non-Frame shift deletion; NSV, non-synonymous variant; CADD phred: Combined Annotation Dependent Depletion; N. PD: Number of PD patients carrying the specific variant; MAF*, highest allelic frequency annotated in public database 1000 Genomes Project (AFR. AMR. EAS. EUR. SAS) and gnomAD (V4.1). **In bold are reported the pathogenic variants identified in the genetic PD cohort (gPD)**.

**Supplementary Table 2.** Demographic and clinical characteristics of male and female PD patients with different PD subtypes.

| Sex | Demographic and clinical information | iPD | | | | gPD | | | | rvPD | | | | p value[a] |
|---|---|---|---|---|---|---|---|---|---|---|---|---|---|---|
| | | N | Median | IQR | | N | Median | IQR | | N | Median | IQR | | |
| Male | Age (Years) | 65 | 67 | 61 | 74 | 64 | 68 | 62 | 72 | 44 | 67 | 61 | 74 | 0.988 |
| | Age at onset (Years) | 65 | 61 | 55 | 66 | 64 | 60 | 56 | 66 | 44 | 60 | 54 | 66 | 0.796 |
| | Years of disease (Years) | 65 | 5 | 3 | 8 | 64 | 6 | 3 | 10 | 44 | 5 | 4 | 10 | 0.766 |
| | LEDD at interview (mg/die) | 65 | 400 | 300 | 560 | 64 | 451 | 320 | 724 | 44 | 500 | 300 | 699 | 0.237 |
| | MDS-UPDRS III | 65 | 24 | 13 | 35 | 64 | 23 | 15 | 31 | 44 | 26 | 15 | 35 | 0.688 |
| Female | Age (Years) | 56 | 70 | 63 | 74 | 60 | 68 | 64 | 74 | 47 | 69 | 60 | 72 | 0.501 |
| | Age at onset (Years) | 56 | 64 | 57 | 69 | 60 | 62 | 53 | 65 | 47 | 60 | 53 | 66 | 0.111 |
| | Years of disease (Years) | 56 | 5 | 3 | 7 | 60 | 7 | 4 | 12 | 47 | 5 | 3 | 10 | **0.033** |
| | LEDD at interview (mg/die) | 56 | 400 | 303 | 600 | 60 | 483 | 300 | 645 | 47 | 410 | 300 | 705 | 0.957 |
| | MDS-UPDRS III | 56 | 21 | 13 | 29 | 60 | 18 | 12 | 30 | 47 | 26 | 16 | 32 | 0.187 |

Abbreviations: iPD, idiopathic Parkinson's disease; gPD, Parkinson's disease with pathogenic variants; rvPD, Parkinson's disease wirh rare variants; N, number of subjects; IQR, interquartile range, LEDD, Levodopa equivalent daily dose; MDS-UPDRS III, Movement Disorders Society Unified Parkinson’s Disease Rating Scale, part III.

[a] Kruskal-Wallis analysis. The significant p value related to disease duration was not confirmed after post-hoc analysis ( p = 0.067).

**Supplementary Table 3.** Robust Volcano plot results related to rvPD compared to HC without sex stratification.

| ***Metabolites*** | ***FC*** | ***p value*** |
|---|---|---|
| L-Glutamic acid | 0.17 | 1.00-07 |
| L-Glutamine | 5.65 | 3.00E-04 |
| Pyruvic acid | 0.25 | 4.00E-02 |

Results related to univariate analysis were conducted using a Robust Volcano plot on concentrations of metabolites detected in the 1 d-CPMG NMR spectrum. The analysis is based on the combination of the Fold change (FC) test, calculated as the ratio of the average concentrations between the rvPD/HC cluster—whose threshold was set to 2—and the p-value derived from the T-Test, considered significant for p-value < 0.05.

**Supplementary Table 4.** Biochemical pathways were identified through Pathway Enrichment analysis in PD patients with rare mutations and HC.

| ***Dysregulated pathways*** | ***Hits*** | ***Raw p*** | ***Holm p*** | ***FDR*** | ***Metabolites*** |
|---|---|---|---|---|---|
| Alanine Metabolism | 4 | 1.57E-06 | 1.00E-03 | 1.57E-02 | Glycine, L-Glutamic acid, L-Alanine, Pyruvic acid. |
| Nicotinate and Nicotinamide Metabolism | 2 | 1.87E-06 | 1.29E-05 | 6.55E-04 | L-Glutamic acid, L-Glutamine. |
| Glucose-Alanine Cycle | 4 | 2.00E-06 | 1.24E-02 | 1.56E-02 | Glucose, L-Glutamic acid, L-Alanine, Pyruvic acid. |
| Warburg Effect | 7 | 2.17E-06 | 1.41E-04 | 2.54E-02 | Citric acid, Glucose, L-Glutamic acid, Lactic acid, Pyruvic acid, Succinic acid, L-Glutamine. |
| Cysteine Metabolism | 2 | 4.51E-06 | 2.84E-03 | 3.94E-02 | L-Glutamic acid, Pyruvic acid. |
| Amino Sugar Metabolism | 4 | 5.30E-06 | 3.71E-06 | 3.71E-06 | Glycine, L-Glutamic acid, Alanine, Pyruvic acid. |
| Urea Cycle | 8 | 6.00E-06 | 3.96E-05 | 8.40E-02 | L-Glutamic acid, L-Glutamine, L-Ornithine, Pyruvic acid, Arginine, Urea, L-Alanine, Aspartate. |
| Aspartate Metabolism | 6 | 8.32E-06 | 5.66E-05 | 1.94E-02 | Acetic acid, L-Glutamic acid, L-Asparagine, Aspartate, L-Arginine, L-Glutamine |
| Glycine and Serine Metabolism | 11 | 2.43E-05 | 1.46E-02 | 1.55E-02 | Betaine, Creatine, Glycine, L-Glutamic acid, L-Alanine, L-Threonine, L-Serine, L-Ornithine, Pyruvic acid, L-Methionine, L-Arginine |
| Ammonia Recycling | 8 | 3.81E-05 | 2.55E-05 | 6.67E-02 | Glycine, L-Glutamic acid, L-Asparagine, L-Histidine, L-Serine, Aspartate, Pyruvic acid, L-Glutamine |
| Glutamate Metabolism | 7 | 4.15E-05 | 2.53E-03 | 2.91E-02 | L-Glutamic acid, L-Alanine, Aspartate, Glycine, Pyruvic acid, Succinic acid, L-Glutamine. |
| Glutathione Metabolism | 3 | 9.31E-05 | 5.49E-02 | 5.43E-03 | Glycine, L-Glutamic acid, L-Alanine. |
| Phenylalanine and Tyrosine Metabolism | 4 | 1.28E-04 | 7.32E-03 | 6.42E-03 | L-Phenylalanine, Tyrosine, Acetoacetate, L-Glutamic acid. |
| Propanoate Metabolism | 3 | 1.10E-03 | 6.40E-03 | 5.94E-03 | 2-Hydroxybutyrate, L-Glutamic acid, Valine. |
| Purine Metabolism | 5 | 1.72E-03 | 9.45E-03 | 7.52E-03 | Glycine, L-Glutamic acid, L-Glutamine,Hypoxanthine, Aspartate |
| Folate Metabolism | 2 | 4.96E-03 | 2.68E-03 | 1.93E-02 | Formic acid, Glutamic acid. |
| Tryptophan Metabolism | 4 | 4.96E-03 | 2.68E-03 | 1.93E-02 | Formic acid, Glutamic acid, Alanine, Tryptophan. |

| | | | | | |
|---|---|---|---|---|---|
| Tyrosine Metabolism | 4 | 1.87E-02 | 1.05E-02 | 4.74E-02 | L-Phenylalanine, Tyrosine, Acetoacetate, L-Glutamic acid. |

Hits are the matched number of metabolites from the user's uploaded data. p-value (Raw p) is calculated from the enriched analysis. The p-value was adjusted according to the Holm-Bonferroni test, considering the number of samples. The false discovery rate (FDR) is the portion of false positives above the user-specified score threshold. Metabolites are the compounds corresponding to Hits.

**Supplementary Table 5.** Robust Volcano plot results related to rvPD compared to HC stratified by sex

| ***rvPD_M vs HC_M*** | ***FC*** | ***p value*** |
|---|---|---|
| L-Glutamic acid | 0.17 | 1.32E-13 |
| Pyruvic acid | 0.17 | 1.19E-04 |
| Acetic acid | 0.25 | 1.43E-03 |
| L-Tryptophan | 6.9 | 5.05E-11 |
| ***rvPD_F vs HC_F*** | ***FC*** | ***p value*** |
| L-Ornithine | 0.20 | 2.51E-11 |
| Pyruvic acid | 0.17 | 3.16E-08 |
| Choline | 0.20 | 1.58E-05 |
| Acetic acid | 0.23 | 3.16E-08 |
| L-Threonine | 4.00 | 1.00E-09 |

Results from univariate analysis were presented using a Robust Volcano plot of metabolite concentrations detected in the 1 d-CPMG NMR spectrum. The analysis combines the Fold change (FC) test, calculated as the ratio of average concentrations between the rvPD_M/HC_M and rvPD_F/HC_F clusters—using a threshold of 2—and the p-value from the T-Test, which is considered significant for $p\text{-value} < 0.05$.

**Supplementary Table 6.** Biochemical pathways were identified through Pathway Enrichment analysis in male and female PD patients with rare mutations compared to sex-matched HC.

| ***Dysregulated pathways: Male*** | ***Hits*** | ***Raw p*** | ***Holm p*** | ***FDR*** | ***Metabolites*** |
|---|---|---|---|---|---|
| Urea Cycle | 8 | 4.77E-10 | 3.34E-08 | 4.77E-10 | L-Glutamic acid, L-Glutamine, L-Ornithine, Pyruvic acid, L-Arginine, Urea, L-Alanine, Aspartate. |
| Glutathione Metabolism | 3 | 1.48E-09 | 1.02E-07 | 1.48E-09 | Glycine, Glutamic acid, Alanine. |
| Glycine and Serine Metabolism | 11 | 2.25E-09 | 1.53E-07 | 2.25E-09 | Betaine, Creatine, Glycine, L-Glutamic acid, L-Alanine, L-Threonine, L-Serine, L-Ornithine, Pyruvic acid, L-Methionine, L-Arginine. |
| Nicotinate and Nicotinamide Metabolism | 2 | 1.44E-08 | 9.65E-07 | 1.44E-08 | L-Glutamic acid, L-Glutamine. |
| Glucose-Alanine Cycle | 4 | 2.11E-07 | 1.39E-05 | 2.11E-07 | Glucose, L-Glutamic acid, L-Alanine, Pyruvic acid. |
| Tryptophan Metabolism | 4 | 2.95E-07 | 1.92E-05 | 2.95E-07 | Formic acid, L-Glutamic acid, L-Alanine, L-Tryptophan. |
| Malate-Aspartate Shuttle | 2 | 3.38E-07 | 2.17E-05 | 3.38E-07 | L-Glutamic acid, Aspartate. |
| Tyrosine Metabolism | 4 | 4.37E-07 | 2.75E-05 | 4.37E-07 | L-Phenylalanine, Tyrosine, Acetoacetate, L-Glutamic acid |
| Alanine Metabolism | 4 | 5.41E-07 | 3.35E-05 | 5.41E-07 | Glycine, L-Glutamic acid, L-Alanine, Pyruvic acid. |
| Warburg Effect | 7 | 1.12E-06 | 6.85E-05 | 1.12E-06 | Citric acid, Glucose, L-Glutamic acid, Lactic acid, Pyruvic acid, Succinic acid, L-Glutamine. |
| Lysine Degradation | 2 | 1.44E-06 | 8.65E-05 | 1.44E-06 | L-Glutamic acid, L-Lysine. |
| Phenylalanine and Tyrosine Metabolism | 4 | 4.74E-06 | 0.00028 | 4.74E-06 | L-Phenylalanine, Tyrosine, Acetoacetate, L-Glutamic acid. |
| Glutamate Metabolism | 7 | 5.50E-06 | 0.000319 | 5.50E-06 | L-Glutamic acid, L-Alanine, Aspartate, Glycine, Pyruvic acid, Succinic acid, L-Glutamine. |
| Aspartate Metabolism | 6 | 3.17E-05 | 0.001809 | 3.17E-05 | Acetic acid, Glutamic acid, Asparagine, Aspartate, L-Arginine, L-Glutamine. |
| Histidine Metabolism | 3 | 4.72E-05 | 0.002641 | 4.72E-05 | L-Methylhistidine, L-Glutamic acid, L-Histidine. |
| ***Dysregulated pathways: Female*** | ***Hits*** | ***Raw p*** | ***Holm p*** | ***FDR*** | ***Metabolites*** |

| | | | | | |
|---|---|---|---|---|---|
| Pyruvic acid Metabolism | 4 | 6.25E-07 | 4.37E-06 | 4.37E-06 | Pyruvic acid, Acetic acid, L-Glutamine, L-Glutamic acid. |
| Glucose-Alanine Cycle | 4 | 8.24E-06 | 4.61E-03 | 3.59E-02 | Glucose, L-Glutamic acid, L-Alanine, Pyruvic acid. |
| Amino Sugar Metabolism | 4 | 9.07E-06 | 6.26E-04 | 3.18E-04 | Pyruvic acid, Acetic acid, Lactic acid,L-Glutamic acid. |
| Gluconeogenesis | 3 | 2.89E-05 | 1.69E-03 | 1.45E-02 | Glucose, Lactic acid, Pyruvic acid,Acetic acid. |
| Fatty Acid Biosynthesis | 3 | 4.20E-04 | 2.77E-02 | 5.42E-03 | Acetic acid, Acetoacetic acid, 3-Hydroxybutyrate. |
| Urea Cycle | 8 | 1.88E-02 | 1.00E-03 | 1.01E-02 | L-Glutamic acid, L-Alanine. Aspartate, L-Ornithine, Pyruvic acid, Urea, L-Arginine, L-Glutamine. |

Hits is the matched number of metabolites from the user uploaded data. p value (Raw p) is calculated from the enriched analysis. The p-value was adjusted according to the Holm-Bonferroni test considering the number of samples. The false discovery rate (FDR) is the portion of false positives above the user-specified score threshold. Metabolites are the compounds corresponding to the Hits.

**Supplementary Table 7.** Biochemical pathways were identified through Pathway Enrichment analysis in male versus female PD patients carrying rare mutations.

| ***Dysregulated pathways: Male vs Female rvPD*** | ***Hits*** | ***Raw p*** | ***Holm p*** | ***FDR*** | ***Metabolites*** |
|---|---|---|---|---|---|
| Selenoamino Acid Metabolism | 2 | 2.37E-03 | 1.66E-02 | 1.66E-02 | L-Alanine; Serine |
| Glucose-Alanine Cycle | 4 | 2.84E-02 | 1.96E-02 | 4.92E-02 | D-Glucose; L-Glutamic acid; L-Alanine; Pyruvic acid |
| Tryptophan Metabolism | 4 | 2.85E-02 | 1.94E-02 | 3.65E-02 | Formic acid; L-Glutamic acid; L-Alanine; Tryptophan |
| Glutathione Metabolism | 3 | 1.60E-02 | 1.08E-02 | 2.81E-02 | Glycine; L-Glutamic acid; L-Alanine |

Hits is the matched number of metabolites from the user-uploaded data. p-value (Raw p) is calculated from the enriched analysis. The p-value was adjusted according to the Holm-Bonferroni test, considering the number of samples. The false discovery rate (FDR) is the portion of false positives above the user-specified score threshold. Metabolites are the compound corresponding to the Hits.

**Supplementary Table 8**. Demographic and clinical characteristics between male and female PD patients carrying rare variants.

| Demographic and clinical information | rvPD | | | | | | | | p value[a] |
|---|---|---|---|---|---|---|---|---|---|
| | Male | | | | Female | | | | |
| | N | Median | IQR | | N | Median | IQR | | |
| Age (Years) | 44 | 67 | 61 | 74 | 47 | 69 | 60 | 72 | 0.886 |
| Age at onset (Years) | 44 | 60 | 54 | 66 | 47 | 60 | 53 | 66 | 0.750 |
| Years of disease (Years) | 44 | 5 | 4 | 10 | 47 | 5 | 3 | 10 | 0.448 |
| LEDD at interview (mg/die) | 44 | 500 | 300 | 699 | 47 | 410 | 300 | 705 | 0.458 |
| MDS-UPDRS III | 44 | 26 | 15 | 35 | 47 | 26 | 16 | 32 | 0.745 |

Abbreviations: rvPD, Parkinson's disease wirh rare variants; N, number of subjects; IQR, interquartile range, LEDD, Levodopa equivalent daily dose; MDS-UPDRS III, Movement Disorders Society Unified Parkinson’s Disease Rating Scale, part III.

[a] Mann Whitney analysis.

**Supplementary Table 9.** Comparison of serum D- and L-amino acid concentrations between PD patients carrying rare variants and healthy controls.

| Amino acids | HC (N=137) | | | rvPD (N=91) | | | Mann-Whitney | ANCOVA[a] | |
|---|---|---|---|---|---|---|---|---|---|
| | Median | IQR | | Median | IQR | | *p* value | $F_{(1;222)}$ | *p* value |
| L-Asp (µM) | 10.9 | 7.7 | 15.6 | 10.7 | 8.2 | 14.6 | 0.514 | | |
| L-Glu (µM) | 27.4 | 18.5 | 41.5 | 24.4 | 16.5 | 34.6 | 0.188 | | |
| L-Asn (µM) | 20.1 | 11.6 | 30.4 | 19.5 | 11.3 | 28.3 | 0.691 | | |
| D-Ser (µM) | 2.7 | 2.1 | 3.1 | 2.6 | 2.1 | 3.1 | 0.518 | | |
| L-Ser (µM) | 72.5 | 43.0 | 106.4 | 62.6 | 43.1 | 97.3 | 0.445 | | |
| D-Ser/total Ser (%) | 3.5 | 2.4 | 5.6 | 3.7 | 2.6 | 5.5 | 0.66 | | |
| L-Gln (µM) | 168.3 | 109.2 | 228.8 | 166.9 | 110 | 223.9 | 0.958 | | |
| Gly (µM) | 141.4 | 104.2 | 196.3 | 141.4 | 101 | 204.5 | 0.97 | | |
| L-Gln/L-Glu | 6.1 | 4.4 | 7.3 | 6.7 | 5.5 | 8.6 | **0.012** | 0.815 | 0.368 |

Abbreviations: HC, healthy controls; rvPD, Parkinson's disease with rare variants; N, number of subjects; IQR, interquartile range. [a] adjusted for age. sex and LEDD.

**Supplementary Table 10.** Correlation of serum D- and L- amino acids levels with clinical characteristics of male and female rvPD patients.

| Sex | Parameters | Spearman's correlation | L-Asp | L-Glu | L-Asn | D-Ser | L-Ser | D-Ser/total Ser | L-Gln | Gly | L-Gln/L-Glu |
|---|---|---|---|---|---|---|---|---|---|---|---|
| Male | Age at onset | Spearman r | 0.036 | 0.174 | 0.038 | 0.114 | 0.033 | 0.007 | 0.039 | 0.054 | -0.260 |
| | | p value | 0.818 | 0.259 | 0.808 | 0.462 | 0.832 | 0.966 | 0.801 | 0.726 | 0.089 |
| | | N | 44 | 44 | 44 | 44 | 44 | 44 | 44 | 44 | 44 |
| | LEDD | Spearman r | 0.126 | 0.102 | 0.236 | 0.098 | 0.162 | -0.104 | 0.161 | 0.001 | 0.136 |
| | | p value | 0.414 | 0.511 | 0.122 | 0.527 | 0.294 | 0.503 | 0.297 | 0.994 | 0.377 |
| | | N | 44 | 44 | 44 | 44 | 44 | 44 | 44 | 44 | 44 |
| | Disease duration | Spearman r | 0.072 | -0.007 | 0.138 | 0.301 | 0.093 | 0.038 | 0.098 | 0.071 | 0.188 |
| | | p value | 0.642 | 0.964 | 0.371 | 0.047 | 0.547 | 0.807 | 0.528 | 0.648 | 0.223 |
| | | N | 44 | 44 | 44 | 44 | 44 | 44 | 44 | 44 | 44 |
| Female | Age at onset | Spearman r | -0.255 | -0.352 | -0.258 | -0.182 | -0.260 | 0.147 | -0.220 | -0.103 | 0.262 |
| | | p value | 0.084 | **0.015** | 0.080 | 0.221 | 0.077 | 0.325 | 0.137 | 0.492 | 0.075 |
| | | N | 47 | 47 | 47 | 47 | 47 | 47 | 47 | 47 | 47 |
| | LEDD | Spearman r | 0.123 | 0.172 | 0.204 | 0.213 | 0.216 | -0.007 | 0.233 | 0.132 | 0.088 |
| | | p value | 0.411 | 0.249 | 0.170 | 0.150 | 0.145 | 0.965 | 0.116 | 0.378 | 0.555 |
| | | N | 47 | 47 | 47 | 47 | 47 | 47 | 47 | 47 | 47 |
| | Disease duration | Spearman r | 0.168 | 0.239 | 0.268 | 0.201 | 0.299 | -0.125 | 0.292 | 0.230 | 0.172 |
| | | p value | 0.259 | 0.105 | 0.068 | 0.176 | 0.041 | 0.403 | 0.047 | 0.120 | 0.247 |
| | | N | 47 | 47 | 47 | 47 | 47 | 47 | 47 | 47 | 47 |

Abbreviations: N, number of subjects; LEDD, Levodopa equivalent daily dose. All p values from Spearman's correlation are shown. For age at onset, significant p values are shown in bold. All significant p values related to disease duration and LEDD were not confirmed after partial correlation analysis adjusted for age alone or for both age and disease duration, respectively.

**Supplementary Table 11.** Association analysis of common variants in *SHMT1*, *SHMT2* and *GCSH* with PD in large case-control cohorts.

| Sex<br>Cohort | Comparison<br>(N) | Gene | chr | Position<br>hg38 | nt<br>change | position | SNP id | test | p | OR |
|---|---|---|---|---|---|---|---|---|---|---|
| **Male+Female**<br>(MNI-PD vs MNI-HC) | PD *vs* HC<br>(804 *vs* 282) | *SHMT1* | 17 | 18335698 | G>A | Intronic | rs2273028 | ADD | NS | NS |
| | | *SHMT2* | 12 | 57233934 | G>A | Intronic | rs34095989 | ADD | NS | NS |
| | | *GCSH* | 16 | 81087862 | A>G | Intronic | rs8177904 | ADD | NS | NS |
| **Male+Female**<br>(PDGC *vs* UK-HC) | PD *vs* HC<br>(4,586 vs 43,989) | *SHMT1* | 17 | 18335698 | G>A | Intronic | rs2273028 | FT | NS | NS |
| | | *SHMT2* | 12 | 57233934 | G>A | Intronic | rs34095989 | FT | NS | NS |
| | | *GCSH* | 16 | 81087862 | A>G | Intronic | rs8177904 | FT | NS | NS |

PD, Parkinson's disease; HC, Healthy Controls from MNI cohort genetic biobank; p, p-value calculated with logistic regression model with Plink software; ADD, additive genetic model; OR, Odds Ratio; FT, Fisher exact Test; NS, not significant. *Significance after Bonferroni correction p=3.3E-04 (150 variants). Genes tested in the association analysis: *SRR, DAO, DAOA, SHMT1, SHMT2, GLDC, AMT, GCSH, PHGDH, GRIN1, GRIN2A, GRIN2B.*

**Supplementary Table 12.** Association analysis of the *SHMT1* rs2273028-A allelic variant in human brain tissues.

| Gene Symbol | Genecode Id | Variant Id | Position Hg38 | P-Value | NES | T-statistic | Tissue |
|---|---|---|---|---|---|---|---|
| SHMT1 | ENSG00000176974 | rs2273028 | 17: 18335698 | 0.00076 | 0.24 | 3.4 | Brain - Amygdala |
| SHMT1 | ENSG00000176974 | rs2273028 | 17: 18335698 | **3.2e-7** | 0.25 | 5.3 | Brain - Anterior cingulate cortex (BA24) |
| SHMT1 | ENSG00000176974 | rs2273028 | 17: 18335698 | **3.1e-8** | 0.28 | 5.7 | Brain - Caudate (basal ganglia) |
| SHMT1 | ENSG00000176974 | rs2273028 | 17: 18335698 | 0.025 | -0.15 | -2.3 | Brain - Cerebellum |
| SHMT1 | ENSG00000176974 | rs2273028 | 17: 18335698 | 0.024 | 0.16 | 3.1 | Brain - Cortex |
| SHMT1 | ENSG00000176974 | rs2273028 | 17: 18335698 | 0.018 | 0.11 | 2.4 | Brain - Frontal Cortex (BA9) |
| SHMT1 | ENSG00000176974 | rs2273028 | 17: 18335698 | 0.083 | 0.090 | 1.7 | Brain - Hippocampus |
| SHMT1 | ENSG00000176974 | rs2273028 | 17: 18335698 | 0.027 | 0.13 | 2.2 | Brain - Hypothalamus |
| SHMT1 | ENSG00000176974 | rs2273028 | 17: 18335698 | **0.000012** | 0.25 | 4.5 | Brain - Nucleus accumbens (basal ganglia) |
| SHMT1 | ENSG00000176974 | rs2273028 | 17: 18335698 | 0.012 | 0.14 | 2.5 | Brain - Putamen (basal ganglia) |
| SHMT1 | ENSG00000176974 | rs2273028 | 17: 18335698 | 0.29 | 0.069 | 1.1 | Brain - Substantia nigra |

Genecode Id, id number of the SHMT1 transcript in Ensembl Genome Browser; Variant id, id of the variant in the Single Nucleotide Polymorphism database; Position Hg38, genomic position on Chromosome assembly HG38; NES, enrichment score normalized.

**Checklist**

**1. Reporting Guidelines**
-STROBE Compliance: This study was designed and reported following the Strengthening the Reporting of Observational Studies in Epidemiology (STROBE) guidelines for case-control studies. **Pag. 26 lines 19-20**

-Metabolomics Reporting: Metabolomic data acquisition and processing comply with the Metabolomics Standards Initiative (MSI) guidelines. **Pag. 26 lines 19-20**

**2. Ethical Compliance & Registration**
**-**Institutional Review Board (IRB): Approved by the IRB of IRCCS Neuromed, Italy (Protocols: N°9/2015, N°19/2020, N°4/2023). **Pag. 26 lines 11-13**

-ClinicalTrials.gov Registration: The study is registered under identifiers NCT02403765, NCT04620980, and NCT05721911. **Pag. 26 lines 13-14**

-Declaration of Helsinki: All clinical investigations were conducted according to the principles of the Declaration of Helsinki. **Pag. 26 lines 15-16**

-Informed Consent: Written informed consent was obtained from all study participants. **Pag. 26 line 16**

**3. Data Stratification & Quality Control**
- Genetic Stratification: Patients were stratified into three distinct groups:
iPD: Idiopathic (no variants in the 37-gene panel). **Pag. 28 lines 9-10**
gPD: Genetic (pathogenic mutations in LRRK2, GBA1, TMEM175, PARK2, PINK1, PARK7). **Pag. 28 lines 21-26**
rvPD: Rare Variant (at least one rare variant of uncertain pathogenicity). **Pag. 28 lines 11-16**

- Sex-Matching: Healthy controls (HC) were sex-matched to the PD cohort to minimize confounding bias in metabolomic analysis. **Pag. 29 lines 1-2**

- Confounder Adjustment: Statistical analyses (ANCOVA and partial correlations) were adjusted for age, disease duration, and L-DOPA Equivalent Daily Dose (LEDD). **Pag. 31 lines 25-26**

**4. Technical Validation**
- NMR Quantitation: Metabolite identification and quantification were performed using Chenomx NMR Suite and confirmed with Bayesil software. **Pag. 7 line 1, Pag. 30 lines 1-3**

- HPLC Validation: Amino acid concentrations were independently validated via UHPLC with precolumn derivatization. **Pag. 31 lines 11-14**

- Genetic Validation: Key genetic associations (e.g., SHMT1 rs2273028) were validated in an independent larger cohort (MNI-rvPD, N=371). **Pag. 15 lines 14-16**

**5. Data Availability & Reproducibility**
- Software used: PLINK2 (Genetics) **Pag.33 lines 11-14**, MetaboAnalyst 6.0 (Metabolomics) **Pag.7 lines 1-3 and Pag. 30 lines 13-15**, Chenomx (NMR) **Pag. 7 line 1, Pag. 30 lines 1-3**.

- Public Databases: Variant pathogenicity was cross-referenced with gnomAD v.4.1.0 **Pag. 28 line 6**, and LOVD v.3.0 **Pag.27 lines 24-25**. Gene expression data were sourced from the GTEx portal **Pag. 33 lines 17-18**.

**1. Study Design & Setting (STROBE Items 4 & 5)**
- Design Type: Defined as a case-control observational study. **Pag.6 line 4**

- Study Periods: Two recruitment windows specified (June 2015–Dec 2017 and June 2021–Dec 2023) **Pag. 25 line 19.**

- Location: Parkinson Centre of the IRCCS INM Neuromed, Italy **Pag 25. Lines 16-19.**

**2. Participant Selection & Eligibility (STROBE Item 6)**
-Case Definition: PD diagnosis based on ≥2 cardinal motor signs (tremor, bradykinesia, rigidity) and positive response to L-DOPA. **Pag. 25 lines 23-25**

-Control Definition: Healthy subjects (HC) negative for PD gene mutations, matched for sex with the PD cohort. **Pag.26 line 26 and Pag.27 lines 1-2.**

-Age Threshold: Inclusion limited to individuals aged ≥40 years to maintain cohort relevance. **Pag.26 lines 25-26**

- Exclusion Criteria: Explicitly listed (pre-existing psychiatric conditions, other neurodegenerative diseases like MS or ALS, dementia, depression, and use of specific psychotropic medications). **Pag. 25 lines 25-26 and Pag. 25 lines 1-3**

**3. Data Sources & Clinical Assessment (STROBE Item 8)**
-Clinical Scale: MDS-UPDRS Part III used for motor symptom severity (assessed during the "ON" period). **Pag. 25 line 7.**

-Ancestry: Confirmed European ancestry for all participants. **Pag. 25 lines 17-18**

-Biobank Origin: Subjects selected from the IRCCS Neuromed/IGB-CNR biobank. **Pag.25 lines 16-17**

**4. Genetic Stratification Framework**
- Panel Composition: Use of a 37-gene panel (10 Mendelian PD genes + 27 risk factor genes). **Pag. 27 lines 10-21.**
- Sequencing Method: Whole Exome Sequencing (WES) data analyzed for rare exonic variants (MAF < 0.01 based on gnomAD v.4.1.0). **Pag.27 lines 5-8.**

- Group Classification:

iPD (Idiopathic): No mutations in the 37-gene panel. **Pag. 28 lines 9-10.**
rvPD (Rare Variant): Carrying at least one rare variant of uncertain pathogenicity. **Pag. 28 lines 21-26.**
gPD (Genetic): Carrying known pathogenic mutations (e.g., LRRK2 G2019S, GBA1). **Pag. 28 lines 21-26.**

**5. Laboratory & Analytical Protocols**
- Serum Handling: Standardized 6-hour fasting collection, 30-min clotting, and -80°C storage. **Pag. 29 line 5-9.**

- NMR Parameters: 600 MHz Bruker spectrometer **Pag.29 lines 17-18**, CPMG pulse sequence **Pag.6 line 24**, water presaturation, and Chenomx/Bayesil software for quantification **Pag. 7 line 1, Pag. 30 lines 1-3**.

- HPLC Parameters: Methanol dilution **Pag.31 line 7**, TCA neutralization **Pag.31 line 9**, precolumn derivatization (OPA/NAC) **Pag.31 lines 10-11**, and C18 reversed-phase column separation **Pag.31 line 12.**

- Standardization: Use of anonymized codes and internal reference signals (TSP for NMR; external standards for HPLC) **Pag. 27 lines 5-7 and Pag. 29 line 17**.

**6. Statistical & Confounding Control (STROBE Items 10 & 12)**
- Sample Size: Acknowledged as determined by biobank availability (no formal a priori calculation). **Pag. 26 line 21**

- Normality Testing: Shapiro-Wilk test and q-q plots used to guide test selection. **Pag. 31 line 20**

- Confounder Adjustment: Use of ANCOVA for age, LEDD, and disease duration; Natural log transformation for non-normal distributions. **Pag. 31 lines 25-26 and Pag. 32 lines 1-2.**

- Multivariate Analysis: PLS-DA with 10-fold internal cross-validation **Pag. 30 lines 16-18** and VIP score ranking **Pag. 30 lines 19-22.**

- Multiple Testing: Bonferroni correction applied to p-values in genetic association tests. **Pag. 33 lines 13-14.**
